# Multiple measures of depression to enhance validity of Major Depressive Disorder in the UK Biobank

**DOI:** 10.1101/2020.09.18.20196451

**Authors:** Kylie P Glanville, Jonathan R I Coleman, David M Howard, Oliver Pain, Ken B Hanscombe, Bradley Jermy, Ryan Arathimos, Christopher Hübel, Gerome Breen, Paul F O’Reilly, Cathryn M Lewis

## Abstract

**Background:** The UK Biobank (UKB) contains data with varying degrees of reliability and completeness for assessing depression. A third of participants completed a Mental Health Questionnaire (MHQ) containing the gold-standard Composite International Diagnostic Interview (CIDI) criteria for assessing mental health disorders.

**Aims:** To investigate whether multiple observations of depression from sources other than the MHQ can enhance the validity of Major Depressive Disorder.

**Methods:** In participants who did not complete the MHQ (n = 325k), we calculated the number of other depression measures endorsed, e.g. from hospital episode statistics and interview data. We compared cases defined this way to CIDI-defined cases for several estimates: the variance explained by polygenic risk scores (PRS), area under the curve attributable to PRS, SNP-based heritability, and genetic correlations with summary statistics from the Psychiatric Genomics Consortium Major Depressive Disorder (PGC MDD) GWAS.

**Results:** The strength of the genetic contribution increased with the number of measures endorsed. For example, SNP-based heritability increased from 7% in cases who endorsed only one measure of depression, to 21% in cases who endorsed four or five measures of depression. The strength of the genetic contribution to cases defined by at least two measures approximated that for CIDI-defined cases. Most genetic correlations between UKB and PGC MDD exceeded 0.7, but there was variability between pairwise comparisons.

**Conclusions:** Multiple measures of depression can serve as a reliable approximation for case-status where the CIDI measure is not available, indicating sample size can be optimised using the entire suite of UKB data.

## Introduction

The emergence of large-scale biobank resources has enabled genetic association studies of complex human traits to be performed with unprecedented sample sizes, and led to novel implication of common genetic variants with psychiatric disorders, including Major Depressive Disorder (MDD)^1^. One of the analytical challenges in using national biobank resources is deciding on an approach to define disorder case and control status using the multiple sources of information available, each having varying degrees of reliability and completeness. The UK Biobank (UKB) contains extensive data items that are relevant to psychiatric phenotyping, ranging from electronic health records to self-reported health data, and questionnaires that rely on retrospective recall of symptoms^2^. The extent to which each source of information accurately classifies cases and controls for a given trait influences any study that is performed, by affecting power and interpretation of effect sizes^3^. In genetic studies of polygenic traits, large sample sizes are a prerequisite for performing a genome-wide association study (GWAS), but investigators must balance phenotypic rigour against sample size, and missing data, where individuals do not meet criteria for either cases or controls. These issues are particularly salient in disorders such as MDD, which encompass a spectrum of symptom severity and within-disorder phenotypic heterogeneity^4^.

The impact of sampling decisions, as they relate to the balance between sample size and misclassification bias, has been demonstrated in the MDD GWAS literature. The CONVERGE study^5^ adopted a strategy to reduce phenotypic heterogeneity by recruiting only patients with recurrent MDD, diagnosed by a health professional, from a population of Han Chinese females. This was the first GWAS to identify and replicate genome-wide significant loci, despite having fewer participants (5,303 cases and 5,337 controls) than the largest MDD GWAS at the time (9,240 cases and 9,519 controls^6^), indicating the advantage of a comparatively homogeneous sample.

Other authors have leveraged minimal phenotyping to increase sample size in MDD GWAS. Using data collected by 23andMe, Inc., Hyde, et al.^7^ identified 75,607 individuals who reported receiving a clinical diagnosis of depression and 231,747 without a history of depression, and performed a GWAS in which 15 genome-wide significant loci were identified. Leveraging data from the UKB, Howard, et al.^8^ defined ‘broad depression’ as participants who endorsed ever having seen a General Practitioner or Psychiatrist for ‘nerves, anxiety, tension or depression’. This help-seeking phenotype generated a sample of 113,769 cases and 208,811 controls in which 14 genome-wide significant loci were identified. The Psychiatric Genomics Consortium (PGC) leveraged minimal phenotyping by combining samples from 23andMe and a sub-set of the UKB, with clinically ascertained cases to generate a sample of 116,404 cases and 314,990 controls, yielding 44 genome-wide significant loci^1^. A meta-analysis of the latter three GWAS^7^,^8^,^1^ produced a sample size of 246,363 cases and 561,190 controls, revealing 102 genome-wide significant loci^9^.

Although increasing sample sizes have ostensibly increased genetic discovery, some authors have argued that the genetic architecture differs between minimally- and strictly-defined depression phenotypes, and that the former definition may yield associations with variants that are not specific to MDD. Cai, et al.^10^ compared the genetic architecture of depression phenotypes derived from different sources of information in the UKB. The highest SNP-based heritability (*h*^2^_SNP_ = ∼26%) was observed in participants who met criteria for Lifetime Depression according to the Composite International Diagnostic Criteria Short Form^11^ (CIDI-SF) that comprised part of an online Mental Health Questionnaire (MHQ). The observed *h*^2^_SNP_ was comparatively lower in depression phenotypes derived from other sources of information; touchscreen questionnaires used to define Symptom-based depression^12^ (*h*^2^_SNP_ = 19%) and ‘broad depression’ (*h*^2^_SNP_ = 14%), Hospital Episode Statistics coded as ICD-10^13^ diagnoses (*h*^2^_SNP_ = 12%), and nurse interviews used to define self-reported depression (*h*^2^_SNP_ = 11%). Although a high degree of shared genetic liability was observed between these depression phenotypes, pair-wise genetic correlations (*r*_G_) differed significantly from one, suggesting phenotype-specific genetic effects^10^.

One interpretation of these findings is that the MHQ derivation of Lifetime Depression is the gold-standard for depression phenotyping in the UKB, compared to the other sources of phenotypic data available. However, the MHQ was only completed by a sub-set of 157,366 UKB participants. It is unclear whether repeated endorsement of depression, from sources other than the MHQ, can be used to reduce misclassification in participants who did not complete the MHQ, and thereby increase the sample size of credible depression cases.

Here, we establish five depression measures available in all UKB participants and create case groups determined by the number of depression measures endorsed by individuals who did not complete the MHQ. We observe the strength of the genetic contribution to each case group by estimating the variance in depression liability explained by polygenic risk scores (PRS), area under the curve (AUC) attributable to PRS, and SNP-based heritability. We compare the strength of the genetic contribution in cases determined by number of endorsements with MHQ-derived Lifetime Depression cases to assess whether sample size can be optimised using all available phenotyping sources, without substantially increasing misclassification bias. The choice of a control group also influences effect size estimates in genetic studies^14^, and we additionally explore the use of partially-screened or screened controls. We anticipate that our approach will encourage researchers to consider the benefit of using multiple phenotype sources to aid classifying cases and controls, not just for depression, but for the extensive range of complex human disorders available in the UKB.

## Methods

### Participants and phenotyping

The UKB is a prospective health study of over 500,000 individuals located across the United Kingdom. Participants were aged between 40 and 69 at recruitment (2006-2010) and attended a baseline assessment where information on health was collected with a touchscreen questionnaire and verbal interview^2^. Subsets of participants completed repeat assessments: instance 1) n = 20,335 between 2012-2013; instance 2) n = 42,961 (interview) and n = 48,340 (touchscreen) in 2014; and instance 3) n = 2,843 (interview) and n = 3,081 (touchscreen) in 2019. Participants with valid email addresses (n = 339,092) were invited to complete the online MHQ in 2017^15^. The UKB received ethical approval from the North West - Haydock Research Ethics Committee (reference 16/NW/0274). This study was conducted under application number 18177. Participants provided electronic signed consent at recruitment^2^.

We identified six measures for depression phenotyping (summarised below) and tabulated the number of individuals who met the criteria for each. Full definitions and UKB field codes are given in Supplementary Materials.

### Help-seeking

‘Help-seeking’ cases endorsed either of the following questions at baseline or instance 1 or 2: “Have you ever seen a general practitioner (GP) for nerves, anxiety, tension or depression?”, and “Have you ever seen a psychiatrist for nerves, anxiety, tension or depression?”.

### Self-reported Depression

‘Self-reported Depression’ cases endorsed having experienced depression (past or present) during the verbal interview at baseline or instance 1 or 2.

### Antidepressant Usage

‘Antidepressant Usage’ cases endorsed currently taking antidepressant medications during the verbal interview at baseline or instance 1 or 2.

### Depression (Smith)

At baseline, 172,751 participants completed an extended touchscreen questionnaire which was enriched for psychosocial questions in additional to the help-seeking question. From these data, Smith, et al. (2013)^12^ defined three depression phenotypes, all of which required endorsement for ‘Help-seeking’: 1) Single episode of probable Major Depression, 2) Probable recurrent Major Depression (moderate), and 3) Probable recurrent Major Depression (severe). We refer to these individuals who endorsed ‘Help-seeking’ and also met the additional criteria defined by Smith, et al.^12^ as ‘Depression (Smith)’.

### Hospital (ICD-10)

Hospital Episode Statistics contain diagnoses recorded with the International Classification of Diseases, 10^th^ Revision (ICD-10)^13^. We accessed the UKB Data Portal Record Repository to identify ICD-10 diagnoses recorded between April 1997 to October 2016. ‘Hospital (ICD-10)’ cases were individuals assigned a primary or secondary diagnosis for Depressive episode (F32-F32.9) or Recurrent depressive disorder (F33-F33.9).

### Lifetime Depression (MHQ)

157,366 participants completed the MHQ. We identified individuals with a lifetime history of depression from responses to the CIDI depression module^11^. We adopted scoring criteria previously defined^15^, which is equivalent to the Diagnostic and Statistical Manual of Mental Disorders (DSM) criteria for Major Depressive Disorder^16^. We classified ‘Lifetime Depression (MHQ)’ cases as individuals meeting those criteria.

### Screening

We defined five potential psychosis phenotypes: ‘Self-reported Psychosis’, ‘Antipsychotic Usage’, ‘Bipolar (Smith)’, ‘Hospital (ICD-10) Psychosis’, and ‘Psychosis (MHQ Screen)’. Individuals meeting the criteria for any psychosis phenotype were excluded from analysis (n = 5,482). The derivation of the psychosis phenotypes is provided in Section 2, Supplementary Materials.

### Depression phenotypes determined by number of observed depression measures

We split the UKB cohort by MHQ participation. In individuals who did not participate in the MHQ, we calculated endorsement for five depression phenotypes (‘Help-seeking’, ‘Self-reported Depression’, ‘Antidepressant Usage’, ‘Depression (Smith)’, or ‘Hospital (ICD-10)’,) to derive five independent depression case groups. These groups are referred to as: ‘One Measure’, ‘Two Measures’, ‘Three Measures’, ‘Four Measures’, and ‘Five Measures’. We performed the same exercise in individuals who completed the MHQ to observe the phenotypic correlation between depression measures (excluding the MHQ) in those that met the criteria for Lifetime Depression (MHQ) and those that did not.

### Controls

Two control groups were defined. Controls comprised all UKB participants who did not meet the criteria for any of the depression or psychosis phenotypes. MHQ controls were restricted to those who participated in the MHQ and showed no psychiatric pathology in the MHQ responses. The criteria for controls and MHQ controls is provided in Section 3, Supplementary Materials.

### Genetic quality control (QC)

The UKB performed preliminary QC on genotype data^2^. Using genetic principal components (PCs) provided by the UKB, we performed 4-means clustering on the first two PCs to identify and retain individuals of European ancestry. QC was performed using PLINK v1.9^17^ to remove: variants with missingness > 0.02 (before individual QC), individuals with missingness > 0.02, sex-discordant observations, variants with missingness > 0.02 (after individual QC), variants departing from Hardy-Weinberg Equilibrium (p < 1 x 10^−8^), and variants with minor allele frequency (MAF) < 0.01. Relatedness kinship estimates provided by the UKB were used to identify related pairs (KING r^2^ > 0.044)^18^ and the GreedyRelated^19^ algorithm was used to remove one individual from each pair. FlashPCA2^20^ was used to generate PCs for the European-ancestry subset. The UKB imputed genotype data to the Haplotype Reference Consortium^21^ and the UK10K Consortium^22^ using the IMPUTE4 software^2^. We removed imputed variants with INFO score < 0.4 and/or MAF < 0.01.

### Statistical analyses

We summarised sociodemographic data taken at baseline assessment: age, sex, socio-economic status (SES), body mass index (BMI), smoking status, and self-reported overall health rating, where participants were asked to rate their overall health on a scale of 1 (excellent) to 4 (poor). We tested for significant differences in sociodemographic variables between cases and controls using Welch Two Sample t-tests in R v3.6.2^23^. To investigate the impact of control sampling, all statistical analyses were performed using controls and MHQ controls.

### Polygenic risk score analyses

The PRSice-2 software^24^ was used to perform PRS analyses. PRS were calculated using summary statistics from the latest PGC MDD GWAS^1^. The PGC MDD GWAS was performed on multiple cohorts with varying phenotyping strategies including self-report (UKB and 23andMe), electronic medical records, and clinical ascertainment. We compared the predictive utility of PRS calculated using summary statistics from (1) the full PGC MDD sample (excluding UKB), and (2) a sub-set of the PGC MDD sample with self-reported cases removed (additionally excluding 23andMe). QC was performed on summary statistics to remove variants within the Major Histocompatibility Complex, and variants in linkage disequilibrium (r^2^ > 0.1) with the lead variant within a 250kb region.

We tested for association between PRS calculated at eight p-value thresholds (*P*_T_; 0.001, 0.05, 0.1, 0.2, 0.3, 0.4, 0.5 and 1.0) and case-control status in each UKB depression phenotype using logistic regressions adjusted for six PCs, genotyping batch, and assessment centre (*n*=128 variables). To control for multiple testing across *P*_T_, ten thousand permutations were performed for each model using linear regression for computational efficiency. We report observed and empirical p-values at the optimal *P*_T_ and the corresponding *R*^*2*^ estimates, transformed to the liability scale using lifetime risk of 15%^1^. To increase sample size, ‘Four Measures’ and ‘Five Measures’ cases were combined in subsequent analyses. The predictive ability of PRS was assessed using AUC with the R pROC package^25^. We compared AUC for the Null model (6 PCs, genotyping batch, and assessment centre on depression phenotypes) with the Full model with PRS at the optimal *P*_*T*_, using DeLong’s test for two correlated ROC curves.

### SNP-based heritability and genetic correlation analyses

To overcome computational limitations when performing GWAS with a large number of covariates (*n*=128), we regressed 6 PCs, genotyping batch, and assessment centre on depression case-control status using logistic regression with the glm function in R v3.6.2^23^. GWASs were performed on residuals for the five depression groups (One, Two, Three, Four & Five Measures, and Lifetime Depression (MHQ)) using both controls sets. GWASs were performed in BGENIE v1.2^2^ and summary statistics were uploaded to FUMA^26^ to create Manhattan and QQ plots.

SNP-based heritabilities were calculated with LD Score Regression (LDSC) v1.0.0^27,28^ using summary statistics excluding variants with INFO scores < 0.9 and pre-computed LD Scores (1000 Genomes European data). SNP-based heritabilities were transformed to the liability scale using lifetime risk of 15%^1^ and, for comparison across a range of population prevalences (1% to 60%), using the transformation proposed by Lee, et al. (2012)^29^ (equation 8).

Genetic correlations (*r*_G_) were estimated using LDSC v1.0.0^27,28^. The *r*_G_ between each UKB depression phenotype and PGC depression phenotype was calculated using summary statistics from both the full PGC MDD sample (excluding UKB, 116,404 cases and 314,990 controls), and the sub-set of the PGC MDD sample with self-reported cases removed (excluding UKB and 23andMe, 45,591 cases and 97,674 controls)^1^.

The study design is summarised in Figure 1.

**Figure 1:**
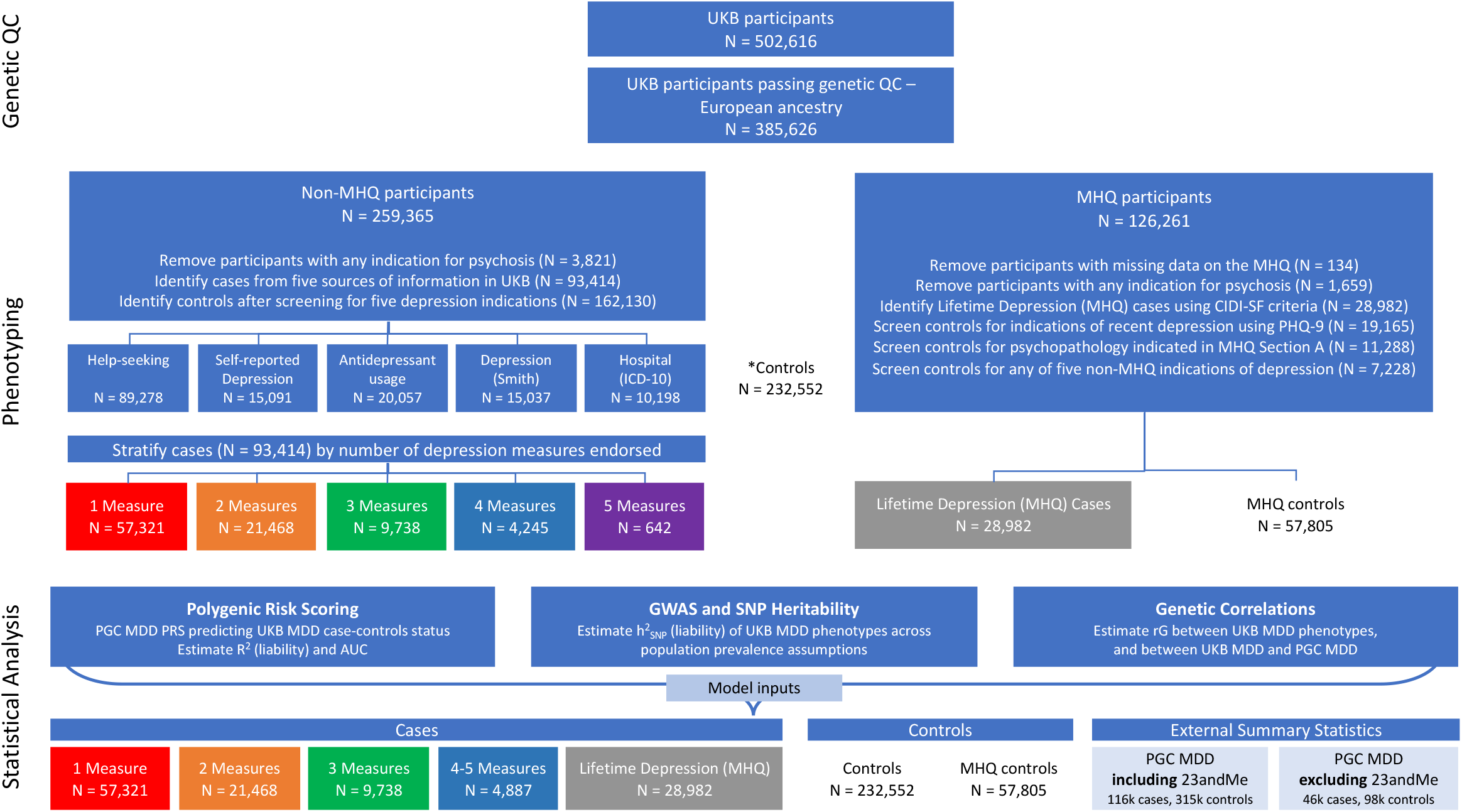
Study design. PHQ-9: Patient Health Questionnaire 9-question version included in the MHQ. Section A, MHQ: participants indicated prior diagnosis for any of sixteen mental health disorders. Refer Supplementary section 3.2 for PHQ-9 and MHQ Section A details. *Controls = UKB participants screened for any of the five psychosis and six depression phenotypes: Help-seeking, Self-reported Depression, Antidepressant Usage, Depression (Smith), Hospital (ICD-10) or Lifetime Depression (MHQ). 162,130 controls are non-MHQ participants. 70,422 controls are MHQ participants (57,805 MHQ controls + 12,617 who didn’t meet CIDI-SF criteria for Lifetime Depression (MHQ) but were excluded from MHQ controls due to psychopathology indicated in MHQ Section A, or above threshold on PHQ-9).

## Results

Of individuals who did not participate in the MHQ, 93,414 met the criteria for at least one other depression phenotype (Table 1). These cases had poorer sociodemographic characteristics than Lifetime Depression (MHQ) cases (N = 28,982) and controls (N = 232,552), including lower SES, higher current smoking prevalence, higher BMI and poorer self-reported heath (all p-values < 1×10^−109^ in pairwise comparisons). The magnitude of difference increased when compared with MHQ controls (N = 57,805), who on average had more favourable sociodemographic outcomes than controls. Lifetime Depression (MHQ) cases also had poorer sociodemographic characteristics compared to both control groups (excluding current smoking status and BMI compared to controls), although the magnitude of case-control differences was attenuated (all p-values < 6×10^−25^ in pairwise comparisons) from that observed with the 93,414 cases derived from sources other than the MHQ. Comparing groups within and outside the MHQ sample, those who participated in the MHQ (N = 126,261) had more favourable sociodemographic characteristics than those who did not participate in the MHQ (N = 259,443), including higher SES, fewer current smokers, lower BMI and higher self-reported heath (all p-values < 3×10^−89^ in pairwise comparisons).

Supplementary Tables 1 to 11 provide the number of participants within sub-categories (e.g. by ICD-10 code) for depression and psychosis in the entire UKB sample.

**Table 1:** Sociodemographic information for depression cases and controls. TDI = Townsend Deprivation Index; negative scores indicate less deprivation. Health Rating was self-reported on a scale of 1 (excellent) to 4 (poor). SD = standard deviation.

|  | Count | Mean age (SD) | Female (%) | TDI (SD) | Current smoker (%) | Mean BMI (SD) | Mean Health Rating (SD) |
| --- | --- | --- | --- | --- | --- | --- | --- |
| Phenotyping in individuals who did not participate in the MHQ |  |  |  |  |  |  |  |
| By source of information |  |  |  |  |  |  |  |
| Hospital (ICD-10) | 10,198 | 56.7 (8.09) | 62% | -0.29 (3.46) | 22% | 29.0 (5.94) | 2.7 (0.83) |
| Self-reported Depression | 15,091 | 55.8 (7.88) | 65% | -0.65 (3.35) | 18% | 28.5 (5.62) | 2.6 (0.82) |
| Depression (Smith) | 15,037 | 56.1 (8.12) | 63% | -1.00 (2.98) | 15% | 28.0 (5.19) | 2.4 (0.77) |
| Antidepressant Usage | 20,057 | 57.0 (7.83) | 68% | -0.70 (3.35) | 18% | 28.9 (5.68) | 2.7 (0.82) |
| Help-seeking | 89,278 | 56.6 (7.97) | 64% | -1.02 (3.20) | 14% | 27.9 (5.16) | 2.3 (0.77) |
| By degree of endorsement |  |  |  |  |  |  |  |
| One Measure | 57,321 | 57 (7.96) | 63% | -1.15 (3.18) | 13% | 27.6 (4.95) | 2.3 (0.74) |
| Two Measures | 21,468 | 57 (8.08) | 64% | -0.92 (3.17) | 15% | 28.0 (5.24) | 2.4 (0.78) |
| Three Measures | 9,738 | 56 (7.90) | 66% | -0.67 (3.35) | 19% | 28.6 (5.60) | 2.6 (0.81) |
| Four Measures | 4,245 | 56 (7.88) | 67% | -0.37 (3.34) | 21% | 29.3 (6.14) | 2.8 (0.84) |
| Five Measures | 642 | 56 (7.63) | 67% | -0.47 (3.23) | 22% | 29.4 (5.64) | 2.8 (0.83) |
| **Total** | **93,414** | **56.7 (7.98)** | **64%** | **-1.01 (3.21)** | **14%** | **27.9 (5.17)** | **2.4 (0.78)** |
| Phenotyping in individuals who participated in the MHQ |  |  |  |  |  |  |  |
| Lifetime Depression (MHQ) | 28,982 | 54.3 (7.53) | 69% | -1.46 (2.95) | 9% | 27.2 (5.04) | 2.1 (0.73) |
| MHQ controls | 57,805 | 56.8 (7.65) | 48% | -2.02 (2.65) | 6% | 26.5 (4.14) | 1.8 (0.62) |
| Phenotyping in all UKB participants (screening for any indication of depression or psychosis) |  |  |  |  |  |  |  |
| Controls | 232,552 | 57.1 (8.10) | 47% | -1.65 (2.89) | 9% | 27.2 (4.53) | 2.0 (0.68) |

Figure 2 shows the 93,414 individuals who did not participate in the MHQ but met the criteria for at least one other depression phenotype, stratified into independent groups according to the number of depression measures endorsed. For each stratum, the number of cases and prevalence as a proportion of controls (N = 232,552) was: One Measure = 57,321 (19.8%), Two Measures = 21,468 (8.5%), Three Measures = 9,738 (4.0%) cases, Four Measures = 4,245 (1.8%), and Five Measures = 642 (0.3%). Of the 28,982 individuals who met CIDI-SF criteria for Lifetime Depression, 9,304 (32%) did not endorse any of the five non-MHQ depression measures and 19,678 (68%) endorsed at least one. Of the 95,486 MHQ participants who did not meet CIDI-SF criteria for Lifetime Depression, 71,848 (75%) did not endorse any of the five non-MHQ depression measures and 23,638 (25%) endorsed at least one. Of individuals who did not meet CIDI-SF criteria, 37,681 (39%) were excluded from MHQ controls for psychopathology indicated within or outside the MHQ as follows: 19,165 excluded for recent depressive symptoms indicated on the PHQ-9 within the MHQ; 11,288 excluded for prior diagnosis of mental health disorders indicated in screening Section A of the MHQ; 7,228 had no indication of psychpathology according to the MHQ but met the criteria for at least one of the five non-MHQ depression measures. The remaining 57,805 participants that did not meet CIDI-SF criteria and endorsed no other measure of depression within or outside the MHQ were defined as MHQ controls. These data are summarised in Figure 3. Supplementary Figure 1 shows the phenotypic agreement between each of the five non-MHQ depression measures within MHQ participants.

**Figure 2:**
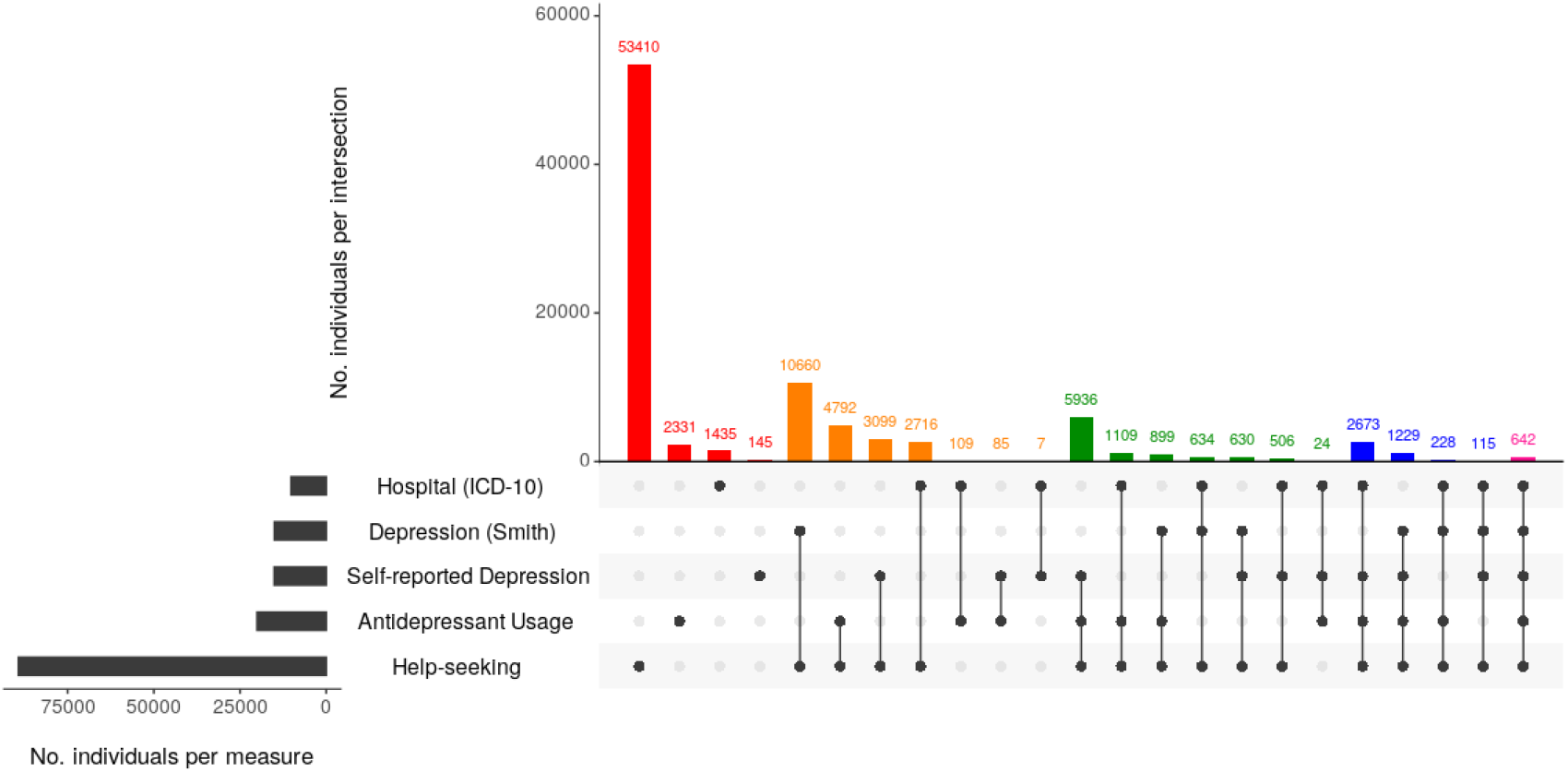
Number of depression measures observed in participants who did not complete the MHQ. Horizontal grey bars indicate the number of individuals who met the criteria for any of the corresponding depression phenotypes. Vertical bars indicate the number of individuals endorsing combinations of the five depression phenotypes. Vertical bars are coloured by the number of depression measures endorsed: red = 1, orange = 2, green = 3, blue = 4 and pink = 5.

**Figure 3:**
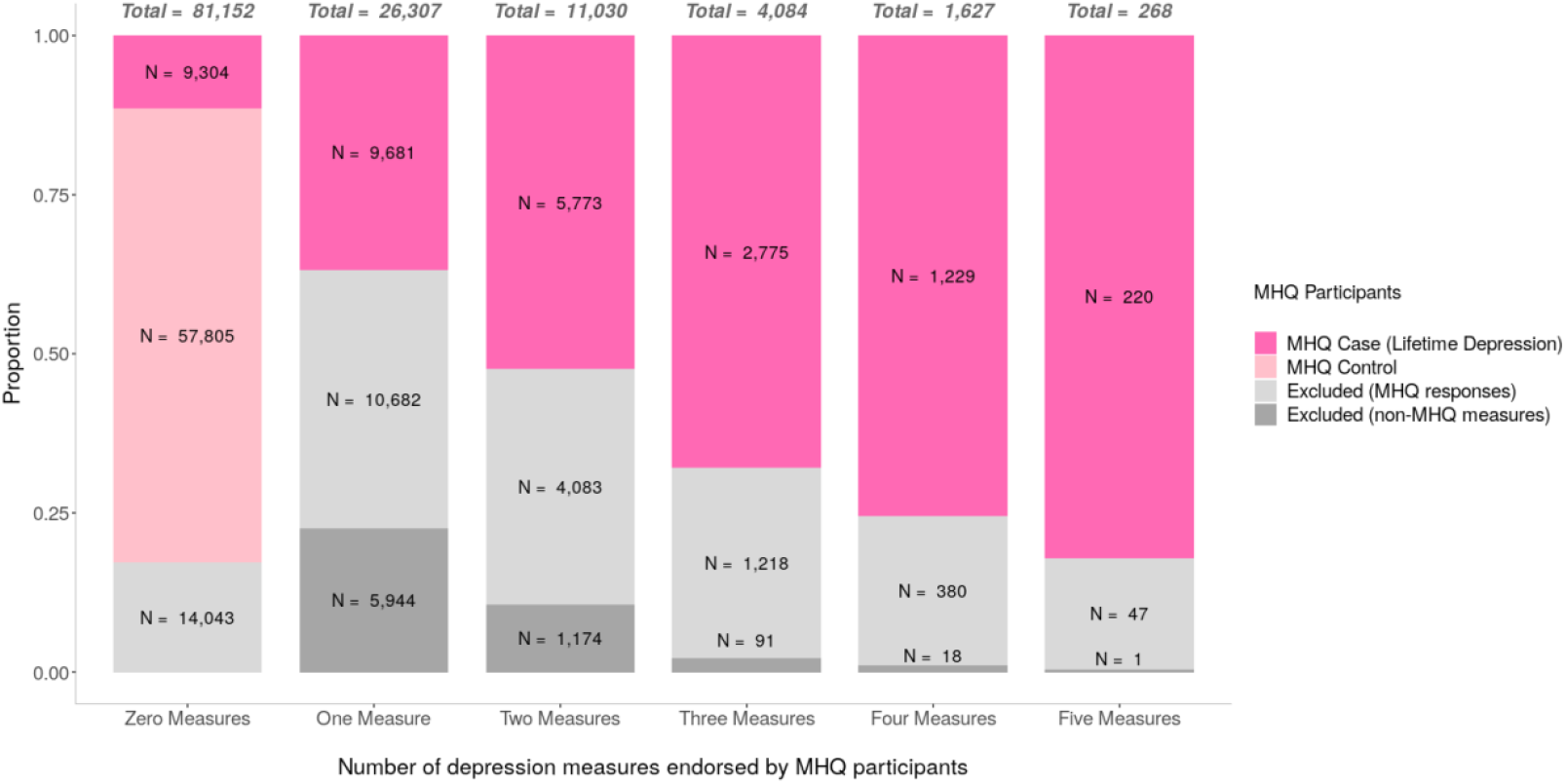
Number of depression measures endorsed by MHQ participants. Categories on the x-axis represent the number of endorsements for the five non-MHQ depression phenotypes (Help-seeking, Self-reported Depression, Antidepressant Usage, Depression (Smith), or Hospital (ICD-10)), with the total number of MHQ participants in each category shown atop each bar. Bars are partitioned by MHQ outcomes. Excluded (MHQ responses) = individuals that did not meet CIDI-SF criteria but had other indications for psychopathology within the MHQ (i.e. PHQ-9 or Screening Section A). Excluded (non-MHQ measures) = individuals that did not meet CIDI-SF criteria and had no indication for psychopathology within the MHQ, but met the criteria for at least one of the five non-MHQ depression measures.

The associations between MDD PRS and case-control status of UKB depression phenotypes were significant (all empirical p-values = 1×10^−4^) (Figure 4). The variance in liability (R^2^) explained by the PRS ranged between 0.52% (One Measure) and 3.54% (Four Measures). Across depression phenotypes, R^2^ increased when cases were compared to MHQ controls, and when PRS were calculated using summary statistics from the full PGC MDD sample (excluding UKB), compared to the sub-set of the PGC MDD (excluding UKB and 23andMe). Full results of each test of association are shown in Supplementary Table 12 and Supplementary Figure 2. Four and Five measures were combined in subsequent analyses to increase power.

**Figure 4:**
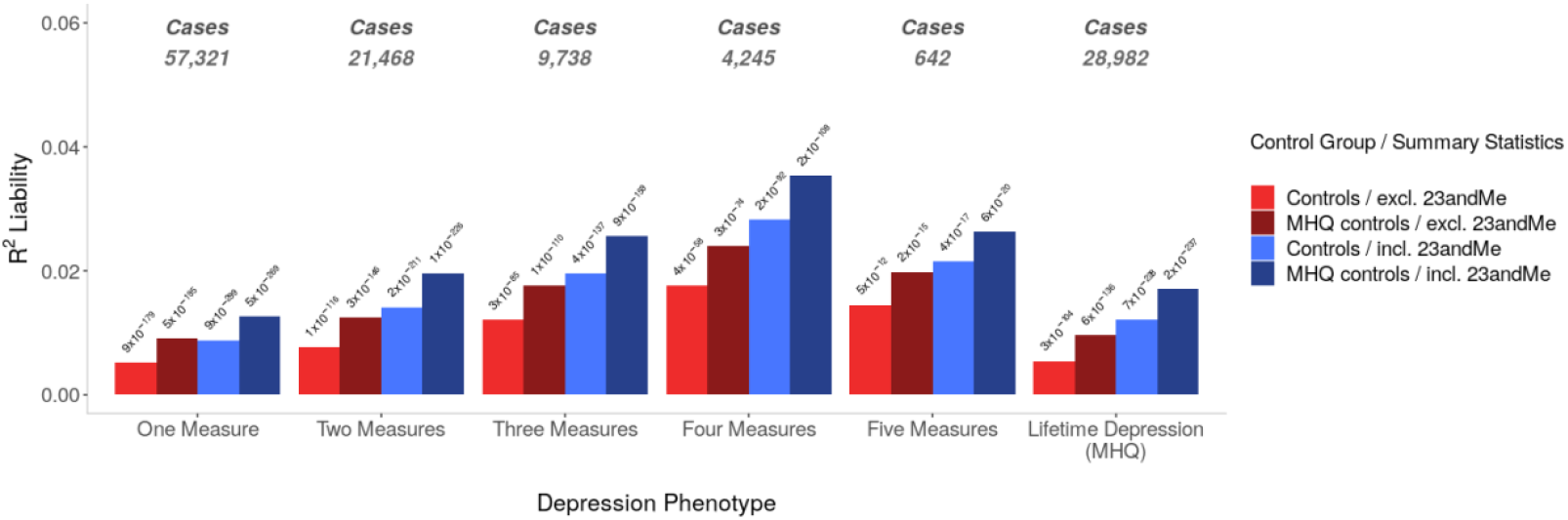
Variances in depression liability explained by PRS. Excl. 23andMe = PRS calculated using summary statistics from the sub-set of the PGC MDD sample (excluding UKB and 23andMe). Incl. 23andMe = PRS calculated using summary statistics from the full PGC MDD sample (excluding UKB). Results are shown for the optimal *P*_T_ for each test of association. R^2^ estimates were transformed to the liability scale using a population prevalence of 15% across all UKB phenotypes. Observed p-values are shown atop each bar.

The differences in AUC between Null and Full Models were significant for each depression phenotype (maximum p-value = 2×10^−25^). The increase in AUC attributable to PRS for models including controls ranged between 1.41% (One Measure) and 3.01% (Three Measures). For models including MHQ controls, the increase in AUC attributable to PRS ranged between 1.29% (One Measure), and 3.60% (Lifetime Depression (MHQ)). AUC attributable to PRS generally increased with the number of depression measures endorsed, maximising in Lifetime Depression (MHQ) when compared to MHQ controls (Figure 5). Supplementary Figure 3 shows ROC curves for Null and Full Models across depression phenotypes.

**Figure 5:**
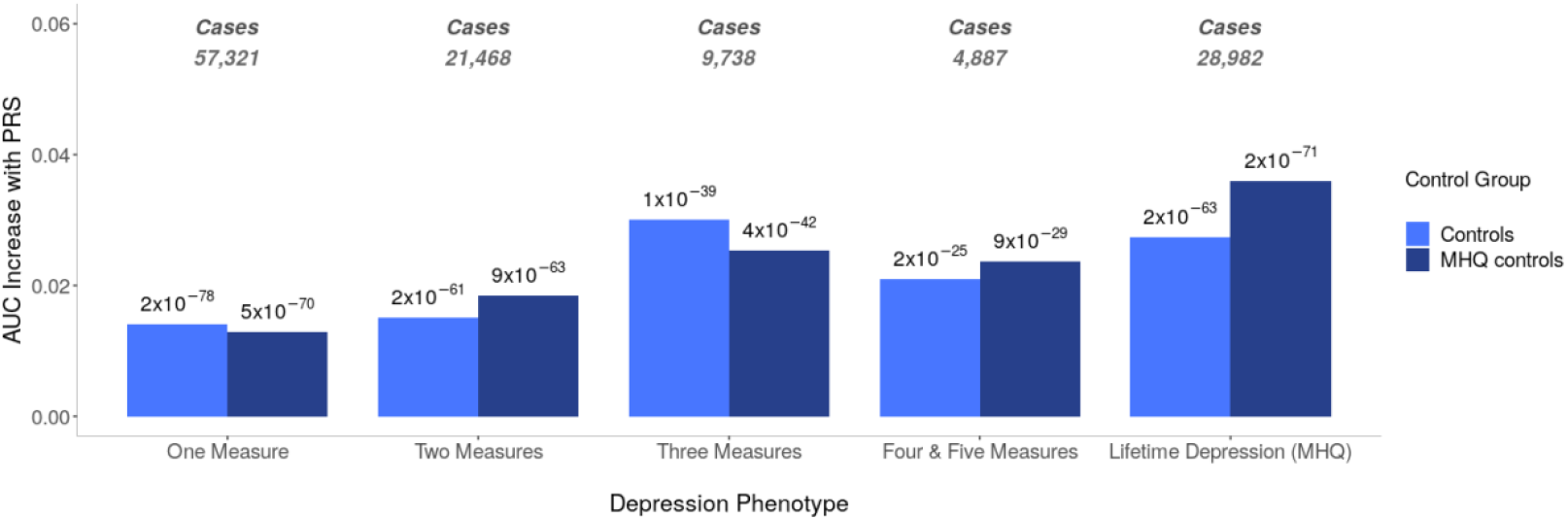
AUC increases attributable to PRS, calculated using full PGC MDD summary statistics (including 23andMe), at the *P*_T_ corresponding to each case-control combination. Y-axis: AUC for Full model minus AUC for Null model. Null vs. Full model p-values estimated with DeLong’s test for two correlated ROC curves are shown atop each bar.

Assuming a population prevalence of 15% across depression phenotypes, SNP-based heritability (*h*^2^_SNP_) estimates ranged between 7% (SE 0.005) in One Measure and 21% (SE 0.029) in Four & Five Measures when GWAS were performed using controls (Figure 6). *h*^2^_SNP_ increased when GWAS were performed with MHQ controls, ranging between 17% (SE 0.009) in One Measure to 33.6% (SE 0.034) in Four & Five Measures. Supplementary Figures 4 to 8 show Manhattan and QQ plots, and Supplementary Tables 14 and 15 show the full results from BGENIE and LDSC.

**Figure 6:**
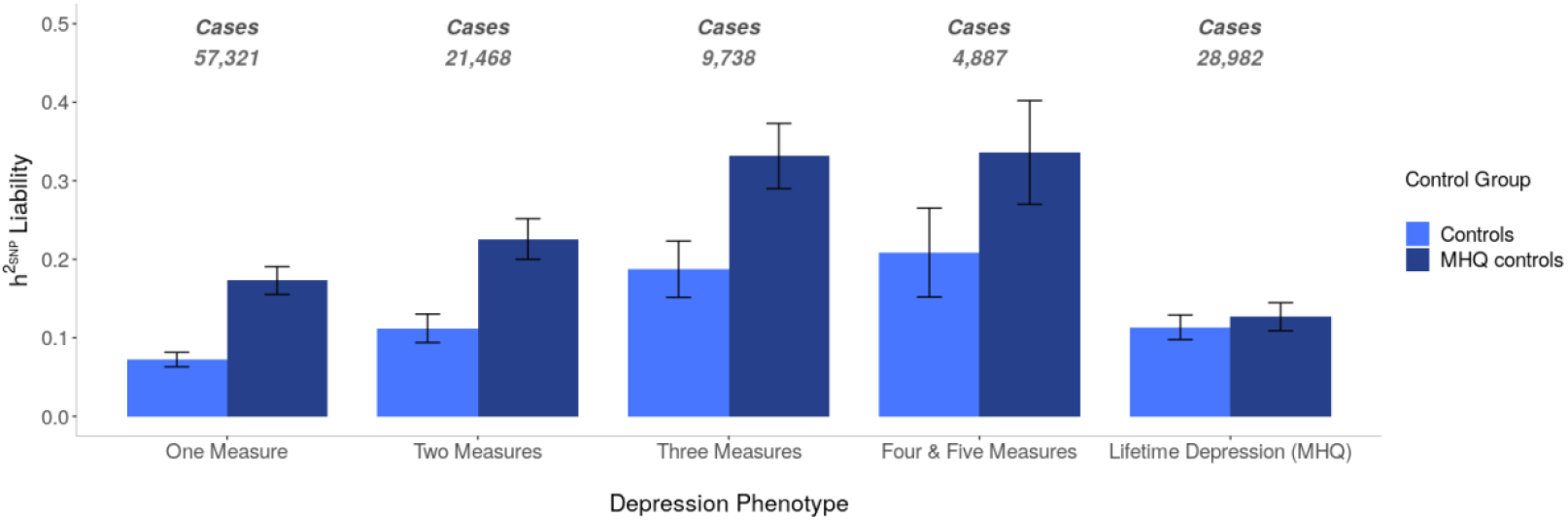
*h*^2^_SNP_ transformed to the liability scale using a population prevalence of 15% across the UKB depression phenotypes on the x-axis. Error bars show 95% confidence intervals.

Across a range of population prevalences between 1% and 60%, higher *h*^2^_SNP_ was observed for models including MHQ controls compared controls (Figure 7). In GWAS using controls, the lowest *h*^2^_SNP_ across the range of population prevalences was in One Measure, followed by Two Measures, Lifetime Depression (MHQ), Three Measures, and Four & Five Measures. We observed near complete overlap in *h*^2^_SNP_ estimates between Two Measures and Lifetime Depression (MHQ), and between the Three Measures and Four & Five Measures. In GWAS using MHQ controls, the lowest *h*^2^_SNP_ across the range of population prevalences was in Lifetime Depression (MHQ), followed by One Measure, Two Measures, Four & Five Measures, and Three Measures. Near complete overlap in *h*^2^_SNP_ estimates was also observed between Three Measures and Four & Five Measures.

**Figure 7:**
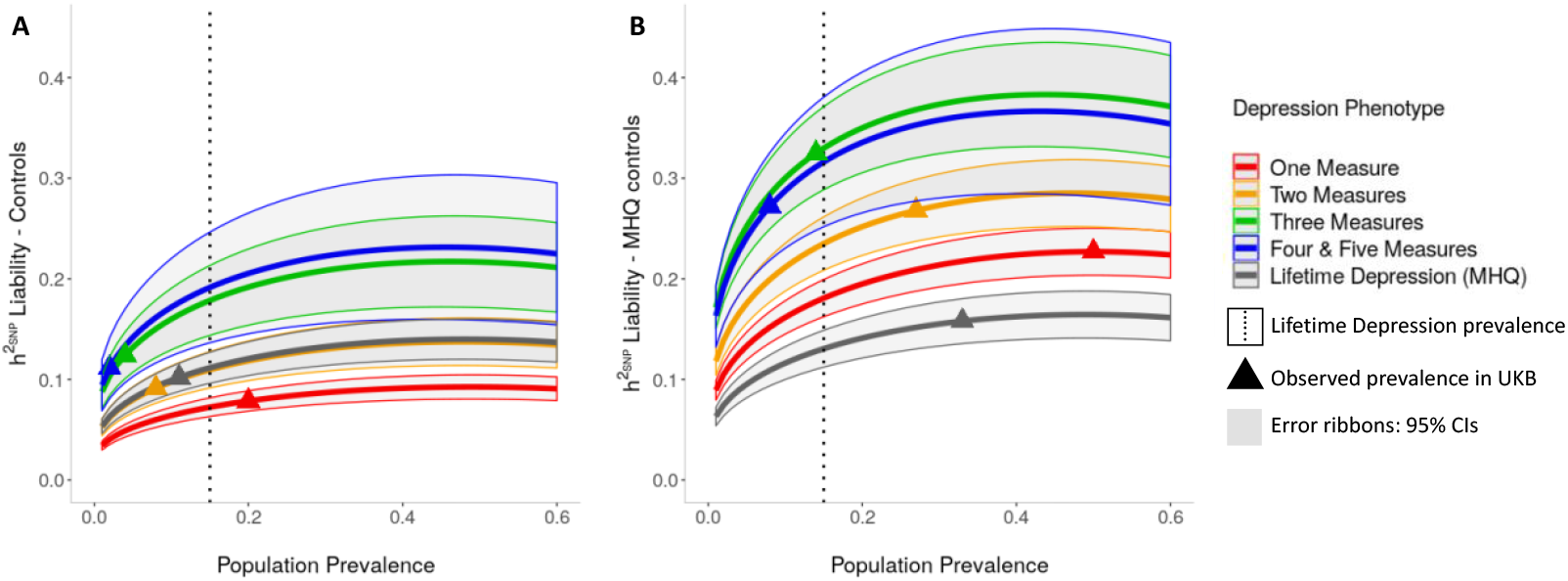
*h*^2^_SNP_ transformed to the liability scale across population prevalence estimates between 1% to 60%. **A**: GWAS performed using controls; **B**: GWAS performed using MHQ controls.

The genetic correlations (*r*_G_) between UKB depression phenotypes and PGC depression phenotypes were between 0.62 and 0.90 (p-value < 6×10^−25^ across all tests for the null hypothesis that *r*_G_ = 0) (Figure 8). The lowest estimate of *r*_G_ was observed between Three Measures (compared to MHQ controls) and the PGC sample including 23andMe (*r*_G_ = 0.62, 95% CI = 0.57-0.67). For the measures of depression, genetic correlations were highest for GWAS using controls, and with summary statistics excluding 23andMe. For Lifetime Depression (MHQ), the highest genetic correlations were for GWAS using MHQ controls, and with the PGC sample excluding 23andMe (r_G_ = 0.90, 95% CI = 0.80-1.00). Supplementary Table 16 and Supplementary Figure 9 show estimates of *r*_G_ between all UKB depression phenotypes.

**Figure 8:**
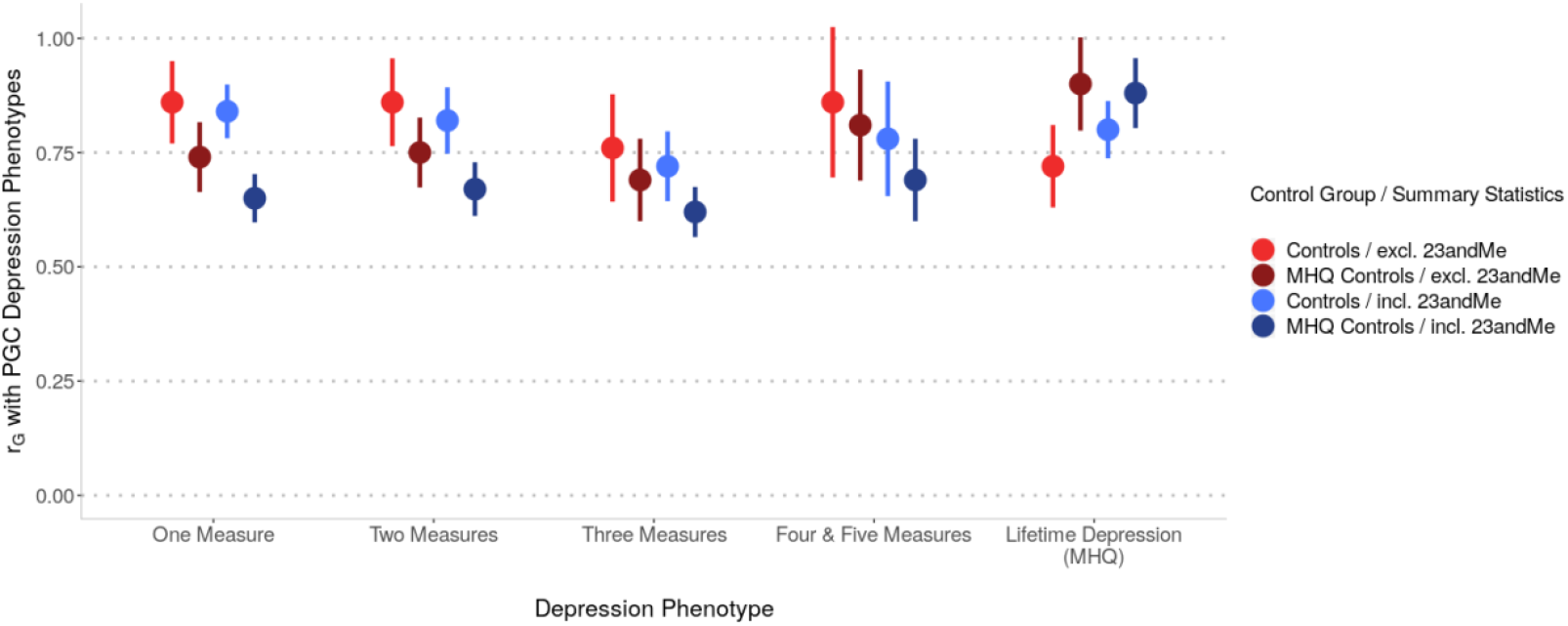
Genetic correlations between the UKB depression phenotypes and PGC depression phenotypes. Excl. 23andMe = red points, summary statistics from the sub-set of the PGC MDD sample (excluding UKB and 23andMe). Incl. 23andMe = blue points, summary statistics from the full PGC MDD sample (excluding UKB). Summary statistics used to estimate *r*_G_ were generated from GWAS of UKB depression phenotypes using controls and MHQ controls. Error bars: 95% Confidence Intervals.

## Discussion

We examined whether multiple endorsements of depression can reduce misclassification and increase the sample of depression cases in the UKB. Our investigation took an approach to classifying depression that aims to fully utilise the UKB by incorporating all sources of information. We found that including at least two measures of depression can serve as a reliable approximation where the MHQ measure is not available and improve case-control classification. Further, increasing the number of measures provides an increasingly reliable approximation.

The results followed from defining independent groups of depression cases according to the number of depression measures endorsed in sources other than the MHQ. We compared cases defined using this approach with CIDI-defined cases for the following: variance explained by PRS, AUC attributable to PRS, and SNP-based heritability. We further explored how these differ using partially-screened controls compared to fully-screened MHQ controls.

Our conclusion is based on three key observations. First, we observed higher values of genetic estimates (variance explained by PRS, AUC attributable to PRS, and SNP-based heritability) with increasing endorsement of depression measures. Second, when cases were defined by two or more measures of depression, these genetic estimates approximated or exceeded those observed in Lifetime Depression (MHQ). Third, control sampling resulted in substantial differences between genetic estimates, which were higher when analyses were performed with MHQ controls.

PRS analyses showed the variance in depression liability increased with the number of measures endorsed, indicating increasing genetic similarity with the PGC MDD sample. The variance explained by PRS was comparable between One Measure and Lifetime Depression (MHQ), although interpretation depends on population prevalence, which is difficult to estimate. By contrast, AUC allows comparisons that are independent of population prevalence. The highest AUC attributable to PRS was observed in Lifetime Depression (MHQ) and was more than double the estimate in One Measure cases. These results indicate that between-group differences in the variance explained by PRS on the liability scale may be masked by equivalent prevalence assumptions across the groups.

However, we found that SNP-based heritability estimates were approximately equivalent for Lifetime Depression (MHQ) and Two Measures across a range of population prevalences between 1% and 60%. Assuming lifetime risk of 15%, *h*^2^_SNP_ for Lifetime Depression (MHQ) ranged between 11-13%, depending on the control group. This range is notably different to the *h*^2^_SNP_ estimate of 26% reported by Cai, et al.^10^ for the corresponding phenotype named ‘LifetimeMDD’. Much of the difference is accounted for by methodology and lifetime risk assumptions. Cai, et al.^10^ used phenotype correlation–genotype correlation (PCGC) software^30^ and the observed prevalence of ‘LifetimeMDD’ in the UKB (24%) to determine liability scale *h*^2^_SNP_. Using LDSC and lifetime risk of 15%, Cai, et al.^10^ report *h*^2^_SNP_ of 16% for ‘LifetimeMDD’, which is modestly higher than our estimate, likely due to minor differences in the derivation of Lifetime Depression (MHQ). Notably, LDSC provides a lower bound of *h*^2^_SNP_ compared to other methods, thus our *h*^2^_SNP_ estimates would increase using other software packages^31^. However, for computational efficiency and consistency with the published literature, we used LDSC and lifetime risk of 15% to calculate *h*^2^_SNP_. Our estimate for Lifetime Depression (MHQ) broadly aligns to the aforementioned GWASs of depression that have adopted the same approach. Using LDSC and lifetime risk of 15%, Hyde, et al.^7^, Howard, et al.^8^, Wray et al.^1^, and Howard, et al.^9^ reported liability *h*^2^_SNP_ of 6%, 10%, 9% and 9% for their respective definitions of depression.

SNP-based heritability increased with the number of measures endorsed, and we posit this results from increasing phenotypic homogeneity within depression case groups. One Measure was comprised mainly of Help-seeking, but also included ostensibly stricter phenotypes including Antidepressant Usage, Hospital (ICD-10) and Self-reported Depression (no participants had a single measure for Depression (Smith) since it requires the endorsement of Help-seeking). However, single observations may reflect indications other than depression. For example, Help-seeking also captures indications for anxiety, and antidepressants can be prescribed for pain management. We therefore regard the number of endorsed measures as more important for phenotypic validity than the specific measure endorsed - those with only one measure are less likely to represent clinical populations than those with multiple measures. The precision of *h*^2^_SNP_ estimates declines in the smaller samples with three or more endorsements, however the confidence internals in these groups showed little or no overlap with cases defined by fewer endorsements or with Lifetime Depression (MHQ), indicating significantly higher *h*^2^_SNP_ across a range of population prevalences. Multiple endorsements may also represent greater severity, but this is not easily demonstrable in the current study because we have not directly measured severity. Cai, et al.^10^ observed higher *h*^2^_SNP_ (32%) in the subset of ‘LifetimeMDD’ who met more stringent criteria for recurrent MDD. Further work is needed to explore disorder severity and SNP-based heritability, which may be possible in the UKB using features such as length of episode and level of impairment.

The pattern of pairwise correlations with the PGC MDD varied across UKB depression phenotypes and was highest with Lifetime Depression (MHQ) (*r*_G_ = 0.9). However, in cases determined by one, two, four or five measures of depression, genetic correlations with the PGC MDD were almost as high, ranging between 0.84 and 0.86. Across UKB depression phenotypes, genetic correlations with the PGC MDD excluding 23andMe were higher than with the PGC MDD including 23andMe. This result indicates greater similarity with the clinically ascertained PGC sample which may lend support to the validity of UKB measures in general.

We observed lower genetic correlations with PGC MDD when GWAS of cases defined by number of endorsements were performed with MHQ controls. Recent work has demonstrated that estimates of genetic parameters increase when sampling controls from the left tail of an underlying liability distribution^14^. We posit that MHQ controls represent the left tail of the liability distribution and this is supported by the observation that MHQ controls were healthier than controls for health indicators correlated with depression prevalence. That is, MHQ controls had higher SES, fewer smokers, lower BMI and better self-reported health ratings than controls. Our results also revealed larger effect sizes across PRS, AUC, and SNP-based heritability analyses when using MHQ controls, compared to controls. MHQ control characteristics may make the UKB dissimilar to the PGC, thus reducing the observed genetic correlation. However, we note that this is not universally supported in the analysis; with Lifetime Depression (MHQ) we observed higher genetic correlations with PGC phenotypes when models included MHQ controls.

Of participants who met CIDI-SF criteria, 32% would have otherwise gone undetected as cases of depression as they did not endorse any of the five non-MHQ measures of depression. Further, of participants who completed the MHQ and did not meet CIDI-SF criteria for Lifetime Depression, 39% were excluded from MHQ controls because they had some other indication for psychopathology within or outside the MHQ, e.g. roughly half were excluded due to recent depressive symptoms indicated on the PHQ-9, but did not fulfil CIDI-SF diagnostic criteria. This is consistent with the view that a percentage of cases would go undiagnosed in primary settings as they never sought help, and a percentage of those who sought help do not fulfill diagnostic criteria for MDD. This highlights the advantage of having both MHQ and non-MHQ sources of information to cross-validate depression phenotypes. Using both sources of information allowed us to define ‘super healthy’ MHQ controls, screened for sub-diagnostic depressive symptoms. While improving the definition of controls may increase power to detect genetic effects, the use of ‘super healthy’ controls omits the intermediate portion of the genetic liability distribution, which can increase SNP-based heritability estimates in the absence of a liability scale correction^14^. We therefore regard the SNP-based heritabilities calculated using controls as the more accurate of the estimates reported here. Future studies with the main objective of genetic discovery may derive power benefits from strict control screening, such as used here to define MHQ controls.

Our results converge on the conclusion that repeated measures of depression may be used to reduce misclassification of depression cases and controls and increase the sample size of credible depression cases in addition to those defined using the MHQ. Cai, et al.^10^ compared depression phenotypes derived from different sources of information in the UKB and showed that the strength of the genetic contribution was highest in CIDI-defined cases. We propose that our findings build upon this work by considering that the number of endorsed measures of depression can be used to decrease misclassification by identifying those participants who perhaps had a single mild episode of depression but would not meet the CIDI diagnostic criteria.

This study enhances the choices available for depression phenotyping in the UKB. The appropriate balance between maximising sample size and minimising misclassification depends naturally on the study to be performed. For genome-wide association studies, two measures showed a high genetic correlation with PGC MDD summary statistics, and individuals with two or more measures would contribute 36,093 cases which could be combined with 28,982 Lifetime Depression (MHQ) cases. Amid increasing use of biobank resources for highly powered psychiatric studies, our study presents a framework that can be adopted for assessing mental health disorders in any biobank that contains multiple sources of information with varying degrees of validity and completeness.

## Limitations

Representativeness is a noted limitation of UKB phenotyping. A healthy volunteer bias has been observed in the UKB^32^, although it has been proposed that this bias does not invalidate exposure-outcome relationships, but may result in attenuated association^33^. However, this selection bias extends to MHQ participation, where we observed more favourable sociodemographic characteristics in MHQ participants compared to non-participants. The differences that we observed in the genetic architecture of depression defined within and outside of the MHQ sample may be influenced by the polygenic basis of MHQ participation, which has been shown to correlate negatively with psychiatric phenotypes^34^. A further limitation of the ability to extrapolate our results is the lack of representation in individuals of diverse ancestries. The literature has demonstrated attenuation in prediction between training and target samples of different ancestry^35^, highlighting the need to build training data in varied ancestral populations.

A further relevant limitation relates to the completeness of the data, and to the opportunity individuals have to endorse specific measures. For example, the extended touchscreen questionnaire used to define Depression (Smith) was only available to approximately a third of the UKB cohort. Regional, procedural or other criteria may have influenced the ability of all measures to be generically applied to the UKB dataset. For instance, recording of data within Scotland excludes linkage to psychiatric hospital episode data. As a result, the reported number of measures may be lower than identified.

Using a simple phenotyping approach, we created independent groups of depression cases determined by the number of depression measures endorsed in the UKB. Our results indicate that two or more endorsements of depression can be used to reduce misclassification between cases and controls, often yielding genetic estimates that approximate, or exceed, the gold-standard CIDI criteria included in the MHQ. While this study has not considered the relative benefit of considering one specific measure over another, the findings of the study highlight that any combination provides a good approximation of depression where the MHQ is not available. With the recent addition of primary care data for approximately half of UKB participants, there is an opportunity to integrate this additional source of information to identify more credible depression cases. We anticipate that this phenotyping approach can be used across other complex traits, to fully utilise the UKB resource.

## Data Availability

Available from UK Biobank subject to standard procedures (www.ukbiobank.ac.uk). The full GWAS summary statistics for the 23andMe discovery data set will be made available through 23andMe to qualified researchers under an agreement with 23andMe that protects the privacy of the 23andMe participants. Please visit https://research.23andme.com/collaborate/#publication for more information and to apply to access the data.

https://research.23andme.com/collaborate/#publication

## Acknowledgements

We thank participants and scientists involved in making the UK Biobank resource available (http://www.ukbiobank.ac.uk/). This study was conducted under UK Biobank application number 18177. We thank the research participants and employees of 23andMe for making this work possible. Statistical analyses were carried out on the King’s Health Partners High Performance Compute Cluster funded with capital equipment grants from the GSTT Charity (TR130505) and Maudsley Charity (980).

## Author contributions

Conceptualisation and study design: KPG, CML, PFO. Analysis and manuscript: KPG. Analytical consultation and interpretation: CML, JRIC, OP, DMH, PFO, BJ. UKB data curation and management: JRIC, GB, RA, KBH, BJ. Genetic data preparation: JRIC. Contributed to data preparation and analysis: CH, OP. Project supervisors: CML, PFO. All authors critically edited the paper.

