## Supplementary for "Multiple measures of depression to enhance validity of Major Depressive Disorder in the UK Biobank"

***Supplement***

### 1 Derivation of depression phenotypes

This section describes the criteria for defining cases for each of the six depression phenotypes derived from different sources of phenotypic information in the UK Biobank. [ID] refers to the corresponding UK Biobank (UKB) Field ID.

#### 1.1 Help-seeking

The criteria for defining ‘Help-seeking’ cases was endorsement of either of the following questions at baseline or the subsequent two repeat assessments:

- "Have you ever seen a general practitioner (GP) for nerves, anxiety, tension or depression?" [ID 2090]; ***OR***
- "Have you ever seen a psychiatrist for nerves, anxiety, tension or depression?" [ID 2100]

#### 1.2 Self-reported Depression

Criteria for defining ‘Self-reported Depression’ cases:

- Endorsed “depression” at baseline or the subsequent two repeat assessments [ID 20002] [UKB Data-Coding 6 = 1286]

#### 1.3 Antidepressant Usage

The criteria for defining ‘Antidepressant Usage’ cases:

- Self-reported taking antidepressant medication at baseline or the subsequent two repeat assessments [ID 20003]. Antidepressant codes [UKB Data-Coding 4]: 1140879616, 1140921600, 1140879540, 1140867878, 1140916282, 1140909806, 1140867888, 1141152732, 1141180212, 1140879634, 1140867876, 1140882236, 1141190158, 1141200564, 1140867726, 1140879620, 1140867818, 1140879630, 1140879628, 1141151946, 1140867948, 1140867624, 1140867756, 1140867884, 1141151978, 1141152736, 1141201834, 1140867690, 1140867640, 1140867920, 1140867850, 1140879544, 1141200570, 1140867934, 1140867758, 1140867914, 1140867820, 1141151982, 1140882244, 1140879556, 1140867852, 1140867860, 1140917460, 1140867938, 1140867856, 1140867922, 1140910820, 1140882312, 1140867944, 1140867784, 1140867812, 1140867668

#### 1.4 Depression (Smith)

The criteria for defining ‘Depression (Smith)’ cases is shown below, comprising the criteria for three depression phenotypes previously defined by Smith, et al. (2013)^1^.

- Single episode of probable Major Depression [ID 20126, Case Coding = 5]
  - ***EITHER***
    - Ever depressed/down for a whole week [ID 4598]; ***AND***
    - At least two weeks duration [ID 4609]; ***AND***
    - Only one episode [ID 4620]; ***AND***
    - Ever seen a GP [ID 2090] ***OR*** a psychiatrist [ID 2100] for nerves, anxiety, depression
  - ***OR***
    - Ever anhedonic (unenthusiasm/uninterest) for a whole week [ID 4631]; ***AND***
    - At least two weeks duration [ID 5375]; ***AND***
    - Only one episode [ID 5386]; ***AND***
    - Ever seen a GP [ID 2090] ***OR*** a psychiatrist [ID 2100] for nerves, anxiety, depression
- Probable recurrent Major Depression (moderate) [ID 20126, Case Coding = 4]
  - ***EITHER***
    - Ever depressed/down for a whole week [ID 4598]; ***AND***
    - At least two weeks duration [ID 4609]; ***AND***
    - At least two episodes [ID 4620]; ***AND***
    - Ever seen a GP [ID 2090] (but not a psychiatrist) for nerves, anxiety, depression
  - ***OR***
    - Ever anhedonic (unenthusiasm/uninterest) for a whole week [ID 4631]; ***AND***
    - At least two weeks duration [ID 5375]; ***AND***
    - At least two episodes [ID 5386]; ***AND***
    - Ever seen a GP [ID 2090] (but not a psychiatrist) for nerves, anxiety, depression
- Probable recurrent Major Depression (severe) [ID 20126, Case Coding = 3]
  - ***EITHER***
    - Ever depressed/down for a whole week [ID 4598]; ***AND***
    - At least two weeks duration [ID 4609]; ***AND***
    - At least two episodes [ID 4620]; ***AND***
    - Ever seen a psychiatrist [ID 2100] for nerves, anxiety, depression
  - ***OR***
    - Ever anhedonic (unenthusiasm/uninterest) for a whole week [ID 4631]; ***AND***
    - At least two weeks duration [ID 5375]; ***AND***
    - At least two episodes [ID 5386]; ***AND***
    - Ever seen a psychiatrist [ID 2100] for nerves, anxiety, depression

#### 1.5 Hospital (ICD-10)

Participants were classified as ‘Hospital (ICD-10)’ cases if they were assigned any of the following primary [ID 41202] or secondary [ID 41204] ICD-10 codes between April 1997 and October 2016:

- Depressive episode: F32, F320, F321, F322, F323, F328, F329.
- Recurrent depressive disorder: F33, F330, F331, F332, F333, F334, F338, F339.

#### 1.6 Lifetime Depression (MHQ)

The criteria for defining ‘Lifetime Depression (MHQ)’ cases is shown below. These criteria mirror those defined by Davis, et al. (2020)^2^ for identifying individuals with a lifetime history of depression (referred to as ‘Depression ever’ in Davis, et al.).

- Endorsed at least one of the two core symptoms:
  - “Have you ever had a time in your life when you felt sad, blue, or depressed for two weeks or more in a row?” [ID 20446]; ***OR***
  - “Have you ever had a time in your life lasting two weeks or more when you lost interest in most things like hobbies, work, or activities that usually give you pleasure?” [ID 20441]; ***AND***
- Endorsed a score above threshold on the following questions:
  - “Please think of the two-week period in your life when your feelings of depression or loss of interest were worst.”
    - “How much of the day did these feelings usually last?” 4 = All day long, 3 = Most of the day, 2 = About half of the day, 1 = Less than half of the day [ID 20436]. Threshold > 2; ***AND***
    - “Did you feel this way” 3 = every day, 2 = almost every day, 1 = less often [ID 20439]. Threshold > 1; ***AND***
    - “Think about your roles at the time of this episode, including study / employment, childcare and housework, leisure pursuits. How much did these problems interfere with your life or activities?” 3 = a lot, 2 = somewhat, 1 = a little [ID 20440]. Threshold > 1; ***AND***
- Endorsed experiencing >=5 symptoms (including core) during worst episode of depression:
  - “Please think of the two-week period in your life when your feelings of depression or loss of interest were worst.”
    - “Did you feel more tired out or low on energy than is usual for you?” [ID 20449]
    - “Did you gain or lose weight without trying, or did you stay about the same weight?” [ID 20536]
    - “Did your sleep change?” [ID 20532]
    - “Did you have a lot more trouble concentrating than usual?” [ID 20435]
    - “People sometimes feel down on themselves, no good, worthless. Did you feel this way?” [ID 20450]
    - “Did you think a lot about death – either your own, someone else’s or death in general?” [ID 20437]

### 2 Derivation of psychosis phenotypes

This section describes the criteria for defining cases for each of the five psychosis phenotypes derived from different sources of phenotypic information in the UK Biobank. Participants who met the criteria for any of the five psychosis phenotypes were excluded from analysis. [ID] refers to the corresponding UK Biobank (UKB) Field ID.

#### 2.1 Self-reported Psychosis

Criteria for defining ‘Self-reported Psychosis’ cases:

- Endorsed “schizophrenia” [UKB Data-Coding 6 = 1289]; ***OR*** “mania/bipolar disorder/manic depression” [UKB Data-Coding 6 = 1291] at baseline or the subsequent two repeat assessments [ID 20002]

#### 2.2 Antipsychotic Usage

The criteria for defining ‘Antipsychotic Usage’ cases:

- Self-reported taking antipsychotic medication at baseline or the subsequent two repeat assessments [ID 20003]. Antipsychotic codes [UKB Data-Coding 4]: 1140868170, 1140928916, 1141152848, 1140867444, 1140879658, 1140868120, 1141153490, 1140867304, 1141152860, 1140867168, 1141195974, 1140867244, 1140867152, 1140909800, 1140867420, 1140879746, 1141177762, 1140867456, 1140867952, 1140867150, 1141167976, 1140882100, 1140867342, 1140863416, 1141202024, 1140882098, 1140867184, 1140867092, 1140882320, 1140910358, 1140867208, 1140909802, 1140867134, 1140867306, 1140867210, 1140867398, 1140867078, 1140867218, 1141201792, 1141200458, 1140867136, 1140879750, 1140867180, 1140867546, 1140928260, 1140927956.

#### 2.3 Bipolar (Smith)

The criteria for defining ‘Bipolar (Smith)’ cases is shown below, comprising the criteria for two bipolar phenotypes previously defined by Smith, et al. (2013)^1^.

- Bipolar Type I (Mania) [ID 20126, Case Coding = 1]
  - Ever manic/hyper 2 days [4642] ***OR*** Ever irritable/argumentative for 2 days [4653]; ***AND***
  - At least 3 from 6156.01 (more active), 6156.02 (more talkative), 6156.03 (needed less sleep), and 6156.04 (more creative/more ideas); ***AND***
  - Duration of a week or more [5663]; ***AND***
  - Needed treatment or caused problems at work [5674]
- Bipolar Type II (Hypomania) [ID 20126, Case Coding = 2]
  - Ever manic/hyper 2 days [4642] ***OR*** Ever irritable/argumentative for 2 days [4653]; ***AND***
  - At least 3 from 6156.01 (more active), 6156.02 (more talkative), 6156.03 (needed less sleep), and 6156.04 (more creative/more ideas); ***AND***
  - Duration of a week or more [5663]

#### 2.4 Hospital (ICD-10) Psychosis

Participants were classified as ‘Hospital (ICD-10) Psychosis’ cases if they were assigned any of the following primary [ID 41202] or secondary [ID 41204] ICD-10 codes between April 1997 and October 2016:

- Schizophrenia, schizotypal and delusional disorders: F20, F200, F201, F202, F203, F204, F205, F206, F208, F209, F21, F22, F220, F228, F229, F23, F230, F231, F232, F233, F238, F239, F24, F25, F250, F251, F252, F258, F259, F28, F29.
- Mood [affective] disorders (excluding Depression codes F32-F33): F30, F300, F301, F302, F308, F309, F31, F310, F311, F312, F313, F314, F315, F316, F317, F318, F319, F34, F340, F341, F348, F349, F38, F380, F381, F388, F39.

#### 2.5 Psychosis (MHQ Screen)

Criteria for defining ‘Psychosis (MHQ Screen)’ cases:

- Endorsed the question (from the MHQ, Section A, screening questions): “Have you been diagnosed with one or more of the following mental health problems by a professional, even if you don’t have it currently?” [ID 20544] for:
  - “Schizophrenia” [UKB Data-Coding 1401 = 2]; ***OR***
  - “Any other type of psychosis or psychotic illness” [UKB Data-Coding 1401 = 3]; ***OR***
  - “Mania, hypomania, bipolar or manic-depression” [UKB Data-Coding 1401 = 10].

### 3 Derivation of Controls

#### 3.1 Controls

Controls comprised all UK Biobank participants who did not meet the criteria for depression or psychosis indications as follows:

- Did not meet the criteria for any indication of depression, i.e.: ‘Lifetime Depression (MHQ)’, ‘Help-seeking’, ‘Depression (Smith)’, ‘Hospital (ICD-10)’, ‘Self-reported Depression’, or ‘Antidepressant Usage’; ***AND***
- Did not meet the criteria for any indication of psychosis, i.e.: ‘Psychosis (MHQ Screen)’, ‘Bipolar (Smith)’, ‘Hospital (ICD-10) Psychosis’, ‘Self-reported Psychosis’, or ‘Antipsychotic Usage’; ***AND***
- Did not endorse the question “Have you been diagnosed with one or more of the following mental health problems by a professional, even if you don’t have it currently?” [ID 20544] for depression [UKB Data-Coding 1401 = 11] in the MHQ, Section A, screening questions.

#### 3.2 MHQ controls

MHQ controls comprised UK Biobank participants who completed the MHQ, met the same criteria as controls, and also met the following criteria:

- Did not endorse the question “Have you been diagnosed with one or more of the following mental health problems by a professional, even if you don’t have it currently?” [ID 20544] for any of the disorders coded 1 to 18 [UKB Data-Coding 1401] in the MHQ, Section A, screening questions. UKB Data-Coding 1401:
  - 1: Social anxiety or social phobia
  - 2: Schizophrenia
  - 3: Any other type of psychosis or psychotic illness
  - 4: A personality disorder
  - 5: Any other phobia
  - 6: Panic attacks
  - 7: Obsessive compulsive disorder
  - 10: Mania hypomania bipolar or manic-depression
  - 11: Depression
  - 12: Bulimia nervosa
  - 13: Psychological over-eating or binge-eating
  - 14: Autism, Aspergers or autistic spectrum disorder
  - 15: Anxiety nerves or generalized anxiety disorder
  - 16: Anorexia nervosa
  - 17: Agoraphobia
  - 18: Attention deficit or attention deficit and hyperactivity disorder; ***AND***
- Scored < 5 on the Patient Health Questionnaire 9-question version (PHQ-9)^3^, from Section B1 of the MHQ:
  - “Over the last 2 weeks, how often have you been bothered by any of the following problems?”
    - “a. Little interest or pleasure in doing things” [ID 20514]
    - “b. Feeling down, depressed, or hopeless” [ID 20510]
    - “c. Trouble falling or staying asleep, or sleeping too much” [ID 20534]
    - “d. Feeling tired or having little energy” [ID 20519]
    - “e. Poor appetite or overeating” [ID 20511]
    - “f. Feeling bad about yourself or that you are a failure or have let yourself or your family down” [ID 20507]
    - “g. Trouble concentrating on things, such as reading the newspaper or watching television” [ID 20508]
    - “h. Moving or speaking so slowly that other people could have noticed? Or the opposite —being so fidgety or restless that you have been moving around a lot more than usual” [ID 20518]
    - “i. Thoughts that you would be better off dead or of hurting yourself in some way” [ID 20513]
      - Scoring for items a. to i. on PHQ9: 4 = nearly every day, 3 = more than half the days, 2 = several days, 1 = not at all.
      - We subtracted 1 from each score and summed across a. to i. Thus, the minimum possible score on the PHQ9 was 0 (e.g. participant responded “not at all” for items a. to i.) and the maximum possible score on the PHQ9 was 27 (e.g. participant responded “nearly every day” for items a. to i.); ***AND***
- Provided sufficient information in Section B (Lifetime Depression) of the MHQ. Participants who responded “prefer not to respond” were excluded from MHQ controls.

### 4 Demographic data

We used the baseline assessment for the following six variables to summarise demographic data in cases and controls.

- Age at recruitment [ID 21002]
- Sex [ID 31]
- Townsend deprivation index [ID 189] – referred to as SES in our study
- Body mass index [ID 21001]
- Smoking status [ID 20116]
  - UKB Data-Coding 90: -3 = Prefer not to answer, 0 = Never, 1 = Previous, 2 = Current
- Overall health rating [ID 2178]. Participants were asked "In general how would you rate your overall health?".
  - UKB Data-Coding 100508: 1 = Excellent, 2 = Good, 3 = Fair, 4 = Poor, -1 = Do not know, -3 = Prefer not to answer
  - We coded negative integers as NA to calculate the average health rating using valid responses

### 5 Count of individuals per phenotypic sub-category

#### 5.1 Depression

Supplementary Tables 1 to 6 show the number of individuals who passed QC and met the criteria for each of the sub-categories of depression phenotypes (before screening for psychosis).

**Supplementary Table 1:** Lifetime Depression (MHQ)

| MHQ | Number |
| --- | --- |
| Lifetime Depression (MHQ) | 29,344 |

**Supplementary Table 2:** Help-seeking

| Help-seeking | Number |
| --- | --- |
| Have you ever seen a GP for nerves, anxiety, tension or depression? | 133,235 |
| Have you ever seen a psychiatrist for nerves, anxiety, tension or depression? | 45,302 |

**Supplementary Table 3:** Depression (Smith)

| Depression (Smith) | Number |
| --- | --- |
| Single episode of probable major depression | 6,154 |
| Probable recurrent major depression (moderate) | 11,654 |
| Probable recurrent major depression (severe) | 6,828 |

**Supplementary Table 4:** Hospital (ICD-10)

| ICD-10 sub-codes | Primary | Secondary |
| --- | --- | --- |
| F32 Depressive episode | | |
| F32 Depressive episode | 3 | 0 |
| F32.0 Mild depressive episode | 60 | 70 |
| F32.1 Moderate depressive episode | 198 | 60 |
| F32.2 Severe depressive episode without psychotic symptoms | 213 | 94 |
| F32.3 Severe depressive episode with psychotic symptoms | 213 | 74 |
| F32.8 Other depressive episodes | 12 | 25 |
| F32.9 Depressive episode, unspecified | 544 | 13,660 |
| F33 Recurrent depressive disorder | | |
| F33.0 Recurrent depressive disorder, current episode mild | 38 | 12 |
| F33.1 Recurrent depressive disorder, current episode moderate | 133 | 18 |
| F33.2 Recurrent depressive disorder, current episode severe without psychotic symptoms | 106 | 30 |
| F33.3 Recurrent depressive disorder, current episode severe with psychotic symptoms | 94 | 13 |
| F33.4 Recurrent depressive disorder, currently in remission | 8 | 7 |
| F33.8 Other recurrent depressive disorders | 2 | 5 |
| F33.9 Recurrent depressive disorder, unspecified | 125 | 338 |

**Supplementary Table 5:** Self-reported Depression

| Self-reported disorder | Number |
| --- | --- |
| Depression | 23,628 |

**Supplementary Table 6:** Antidepressant usage

| Medication | Number |
| --- | --- |
| amitriptyline | 7,458 |
| citalopram | 6,957 |
| fluoxetine | 4,578 |
| sertraline | 1,833 |
| venlafaxine | 1,624 |
| dosulepin | 1,354 |
| paroxetine | 1,301 |
| mirtazapine | 1,121 |
| escitalopram | 711 |
| trazodone | 563 |
| prozac 20mg capsule | 544 |
| seroxat 20mg tablet | 540 |
| cipralex 5mg tablet | 349 |
| duloxetine | 354 |
| lofepramine | 296 |
| clomipramine | 257 |
| nortriptyline | 252 |
| imipramine | 197 |
| dothiepin | 148 |
| cipramil 10mg tablet | 132 |
| amitriptyline hydrochloride+perphenazine 10mg/2mg tablet | 90 |
| prothiaden 25mg capsule | 75 |
| trimipramine | 69 |
| lustral 50mg tablet | 60 |
| reboxetine | 39 |
| zispin 30mg tablet | 50 |
| cymbalta 30mg gastro-resistant capsule | 38 |
| anafranil 10mg capsule | 36 |
| doxepin | 29 |
| moclobemide | 24 |
| phenelzine | 26 |
| fluvoxamine | 27 |
| yentreve 20mg gastro-resistant capsule | 17 |
| triptafen tablet | 23 |
| surmontil 10mg tablet | 17 |
| tranylcypromine | 13 |
| allegron 10mg tablet | 5 |
| edronax 4mg tablet | 6 |
| molipaxin 50mg capsule | 4 |
| mianserin | 3 |
| nardil 15mg tablet | 4 |
| faverin 50mg tablet | 1 |
| nefazodone | 3 |
| amitriptyline+chlordiazepoxide 12.5mg/5mg capsule | 0 |
| isocarboxazid | 0 |
| manerix 150mg tablet | 1 |
| maoi - tranylcypromine | 1 |
| sinequan 10mg capsule | 2 |
| tranylcypromine+trifluoperazine 10mg/1mg tablet | 1 |
| ludiomil 10mg tablet | 1 |
| norval 10mg tablet | 1 |
| tryptizol 10mg tablet | 1 |

#### 5.2 Psychosis

Supplementary Tables 7 to 11 show the number of individuals who passed QC and met the criteria for sub-categories of psychosis phenotypes. These individuals were excluded from analyses.

**Supplementary Table 7:** Psychosis (MHQ screen)

| MHQ screening | Number |
| --- | --- |
| Schizophrenia | 112 |
| Any other type of psychosis or psychotic illness | 483 |
| Mania hypomania bipolar or manic-depression | 649 |

**Supplementary Table 8:** Bipolar (Smith)

| Bipolar (Smith) | Number |
| --- | --- |
| Probable bipolar disorder (type I) | 614 |
| Probable bipolar disorder (type II) | 565 |

**Supplementary Table 9:** Hospital (ICD-10) Psychosis

| ICD-10 sub-codes | Primary | Secondary |
| --- | --- | --- |
| F20-F29 Schizophrenia, schizotypal and delusional disorders | | |
| F20 Schizophrenia | 2 | 1 |
| F20.0 Paranoid schizophrenia | 182 | 114 |
| F20.1 Hebephrenic schizophrenia | 5 | 1 |
| F20.2 Catatonic schizophrenia | 6 | 3 |
| F20.3 Undifferentiated schizophrenia | 4 | 0 |
| F20.4 Postschizophrenic depression | 3 | 0 |
| F20.5 Residual schizophrenia | 9 | 13 |
| F20.6 Simple schizophrenia | 2 | 4 |
| F20.8 Other schizophrenia | 6 | 5 |
| F20.9 Schizophrenia, unspecified | 109 | 422 |
| F21 Schizotypal disorder | 5 | 7 |
| F22.0 Delusional disorder | 83 | 135 |
| F22.8 Other persistent delusional disorders | 2 | 1 |
| F22.9 Persistent delusional disorder, unspecified | 15 | 10 |
| F23 Acute and transient psychotic disorders | 0 | 1 |
| F23.0 Acute polymorphic psychotic disorder without symptoms of schizophrenia | 15 | 1 |
| F23.1 Acute polymorphic psychotic disorder with symptoms of schizophrenia | 8 | 3 |
| F23.2 Acute schizophrenia-like psychotic disorder | 7 | 0 |
| F23.3 Other acute predominantly delusional psychotic disorders | 10 | 4 |
| F23.8 Other acute and transient psychotic disorders | 7 | 1 |
| F23.9 Acute and transient psychotic disorder, unspecified | 76 | 30 |
| F24 Induced delusional disorder | 2 | 0 |
| F25.0 Schizoaffective disorder, manic type | 34 | 7 |
| F25.1 Schizoaffective disorder, depressive type | 26 | 5 |
| F25.2 Schizoaffective disorder, mixed type | 10 | 7 |
| F25.8 Other schizoaffective disorders | 1 | 1 |
| F25.9 Schizoaffective disorder, unspecified | 52 | 68 |
| F28 Other nonorganic psychotic disorders | 2 | 2 |
| F29 Unspecified nonorganic psychosis | 77 | 120 |
| F30-F39 Mood [affective] disorders | | |
| F30.0 Hypomania | 52 | 15 |
| F30.1 Mania without psychotic symptoms | 7 | 0 |
| F30.2 Mania with psychotic symptoms | 30 | 5 |
| F30.8 Other manic episodes | 2 | 1 |
| F30.9 Manic episode, unspecified | 43 | 27 |
| F31.0 Bipolar affective disorder, current episode hypomanic | 150 | 16 |
| F31.1 Bipolar affective disorder, current episode manic without psychotic symptoms | 116 | 13 |
| F31.2 Bipolar affective disorder, current episode manic with psychotic symptoms | 105 | 13 |
| F31.3 Bipolar affective disorder, current episode mild or moderate depression | 84 | 22 |
| F31.4 Bipolar affective disorder, current episode severe depression without psychotic symptoms | 44 | 7 |
| F31.5 Bipolar affective disorder, current episode severe depression with psychotic symptoms | 32 | 3 |
| F31.6 Bipolar affective disorder, current episode mixed | 40 | 9 |
| F31.7 Bipolar affective disorder, currently in remission | 25 | 10 |
| F31.8 Other bipolar affective disorders | 12 | 12 |
| F31.9 Bipolar affective disorder, unspecified | 204 | 828 |
| F34.0 Cyclothymia | 5 | 6 |
| F34.1 Dysthymia | 20 | 40 |
| F34.8 Other persistent mood [affective] disorders | 1 | 1 |
| F34.9 Persistent mood [affective] disorder, unspecified | 0 | 8 |
| F38.0 Other single mood [affective] disorders | 5 | 4 |
| F38.1 Other recurrent mood [affective] disorders | 0 | 1 |
| F38.8 Other specified mood [affective] disorders | 2 | 6 |
| F39 Unspecified mood [affective] disorder | 16 | 37 |

**Supplementary Table 10:** Self-reported Psychosis

| Self-reported disorder | Number |
| --- | --- |
| Schizophrenia | 432 |
| Mania/bipolar disorder/manic depression | 1,110 |

**Supplementary Table 11:** Antipsychotic Usage

| Medication | Number |
| --- | --- |
| prochlorperazine | 551 |
| olanzapine | 431 |
| quetiapine | 236 |
| risperidone | 200 |
| chlorpromazine | 128 |
| trifluoperazine | 78 |
| amisulpride | 77 |
| sulpiride | 55 |
| seroquel 25mg tablet | 60 |
| haloperidol | 47 |
| aripiprazole | 44 |
| stelazine 1mg tablet | 50 |
| depixol 3mg tablet | 44 |
| flupentixol | 51 |
| clozapine | 29 |
| promazine | 29 |
| risperdal 0.5mg tablet | 26 |
| modecate 12.5mg/0.5ml oily injection | 25 |
| fluanxol 500micrograms tablet | 23 |
| flupenthixol | 15 |
| zyprexa 2.5mg tablet | 16 |
| zuclopenthixol | 16 |
| clopixol 2mg tablet | 14 |
| largactil 10mg tablet | 10 |
| abilify 5mg tablet | 8 |
| fluphenazine | 8 |
| haldol 5mg tablet | 5 |
| serenace 500micrograms capsule | 11 |
| clozaril 25mg tablet | 8 |
| cpz - chlorpromazine | 6 |
| perphenazine | 5 |
| levomepromazine | 6 |
| pericyazine | 5 |
| dolmatil 200mg tablet | 3 |
| fentazin 2mg tablet | 3 |
| fluphenazine decanoate | 3 |
| benperidol | 3 |
| pimozide | 3 |
| zaponex 25mg tablet | 1 |
| denzapine 25mg tablet | 1 |
| neulactil 2.5mg tablet | 1 |
| thioridazine | 1 |
| dozic 1mg/ml oral liquid | 1 |
| fluspirilene | 1 |
| panadeine co tablet | 1 |
| sertindole | 0 |

### 6 Number of depression measures observed in MHQ participants

| **A** | **Lifetime Depression (MHQ) cases** |
| --- | --- |
| 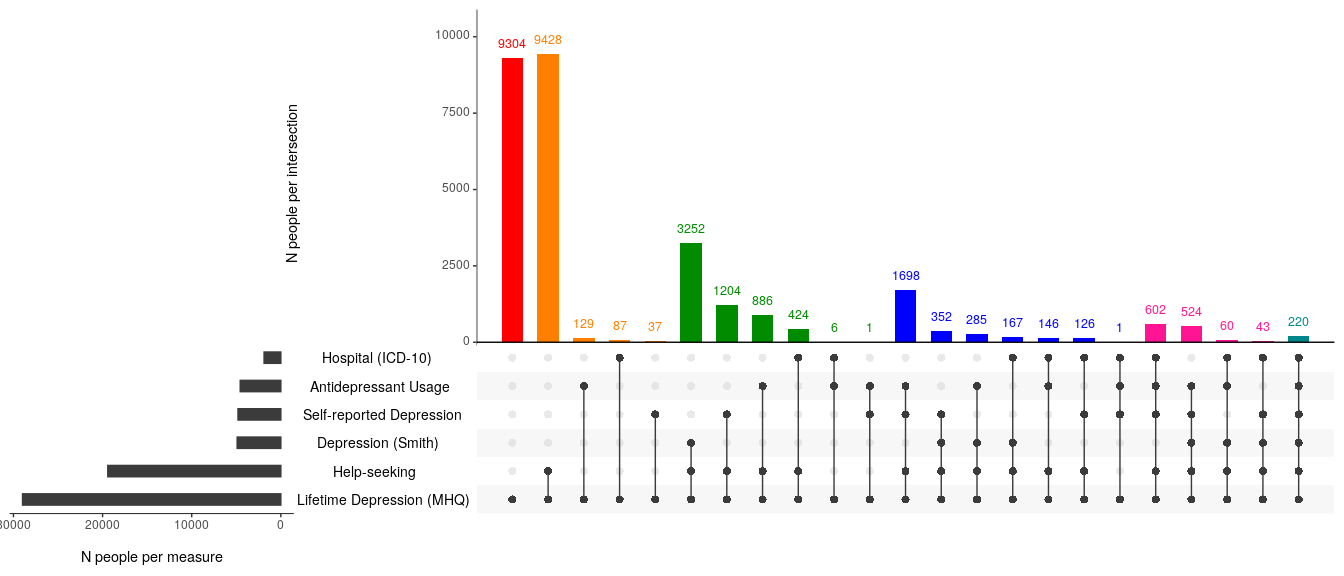 | |
| **B** | **Did not meet CIDI-SF criteria for Lifetime Depression** |
| 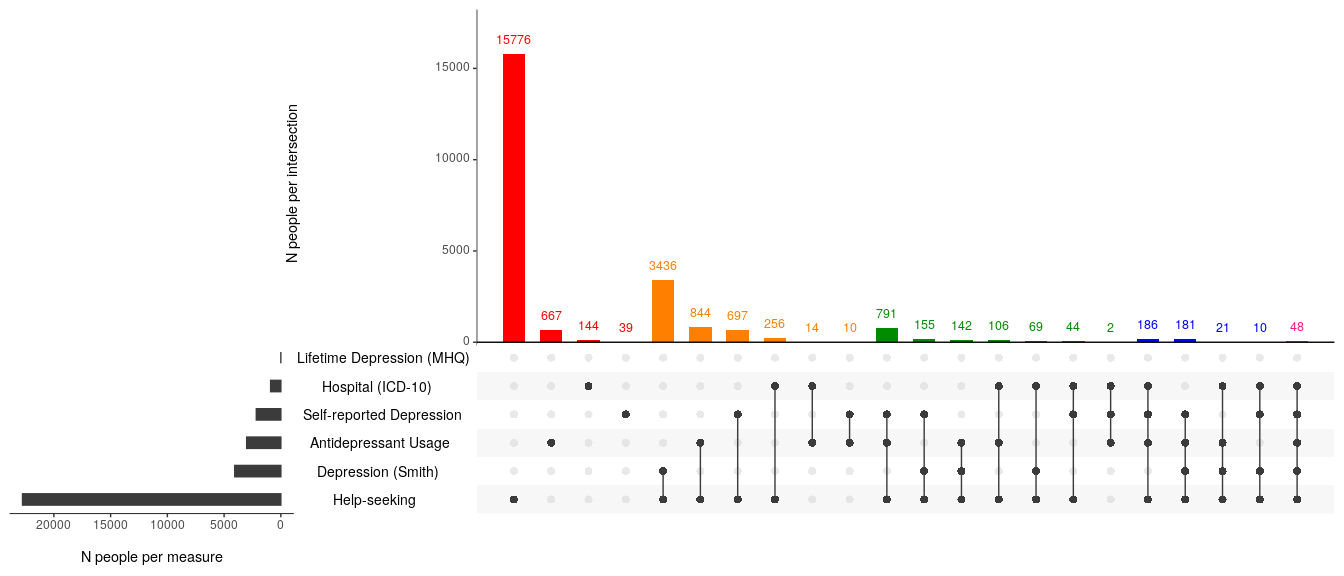 | |

**Supplementary Figure 1**: Number of depression measures observed in MHQ participants. Horizontal grey bars indicate the number of individuals who met the criteria for any of the corresponding depression phenotypes. Vertical bars indicate the number of individuals endorsing combinations of the six depression phenotypes. Vertical bars are coloured by the number of depression measures endorsed: red = 1, orange = 2, green = 3, blue = 4, pink = 5, turquoise = 6. Panel A = MHQ participants who met the criteria for Lifetime Depression (N = 28,982); Panel B = MHQ participants who did not meet the CIDI-SF criteria for Lifetime Depression (Total N = 95,486. N = 37,681 had other indications of depression and were excluded from MHQ controls. N = 57,805 had no other indications of depression and were retained as MHQ controls). N = 1,793 MHQ participants excluded for indications of psychosis or missing data in the MHQ.

### 7 Polygenic risk score analyses results

**Supplementary Table 12**: Results for tests of association between PRS and depression phenotypes**.**

| Base GWAS | Controls | Cases | Optimal p-value threshold (*P*_T_) | No. SNPs used to construct PRS at (*P*_T_) | Population Prevalence | Adjusted Variance explained (Liability R^2^) | Coefficient | Observed p-value | Empirical p-value |
| --- | --- | --- | --- | --- | --- | --- | --- | --- | --- |
| PGC MDD  **Excl**. 23andMe | Controls | One Measure | 0.3 | 45,292 | 0.15 | 0.52% | 0.14 | 9e-179 | 1e-04 |
|  |  | Two Measures | 0.3 | 45,292 | 0.15 | 0.77% | 0.17 | 1e-116 | 1e-04 |
|  |  | Three Measures | 0.3 | 45,292 | 0.15 | 1.21% | 0.21 | 3e-85 | 1e-04 |
|  |  | Four Measures | 0.4 | 55,560 | 0.15 | 1.76% | 0.25 | 4e-58 | 1e-04 |
|  |  | Five Measures | 0.2 | 33,491 | 0.15 | 1.45% | 0.28 | 5e-12 | 1e-04 |
|  |  | Lifetime Depression (MHQ) | 0.5 | 64,483 | 0.15 | 0.54% | 0.14 | 3e-104 | 1e-04 |
| PGC MDD  **Excl**. 23andMe | MHQ controls | One Measure | 0.3 | 45,292 | 0.15 | 0.91% | 0.19 | 5e-195 | 1e-04 |
|  |  | Two Measures | 0.3 | 45,292 | 0.15 | 1.25% | 0.22 | 3e-146 | 1e-04 |
|  |  | Three Measures | 0.3 | 45,292 | 0.15 | 1.77% | 0.25 | 1e-110 | 1e-04 |
|  |  | Four Measures | 0.4 | 55,560 | 0.15 | 2.39% | 0.30 | 3e-74 | 1e-04 |
|  |  | Five Measures | 0.2 | 33,491 | 0.15 | 1.98% | 0.33 | 2e-15 | 1e-04 |
|  |  | Lifetime Depression (MHQ) | 0.3 | 45,292 | 0.15 | 0.96% | 0.18 | 6e-136 | 1e-04 |
| PGC MDD  **Incl**. 23andMe | Controls | One Measure | 0.5 | 67,956 | 0.15 | 0.88% | 0.18 | 9e-299 | 1e-04 |
|  |  | Two Measures | 0.4 | 59,001 | 0.15 | 1.41% | 0.23 | 2e-211 | 1e-04 |
|  |  | Three Measures | 0.2 | 36,975 | 0.15 | 1.96% | 0.26 | 4e-137 | 1e-04 |
|  |  | Four Measures | 0.4 | 59,001 | 0.15 | 2.83% | 0.32 | 2e-92 | 1e-04 |
|  |  | Five Measures | 0.1 | 22,831 | 0.15 | 2.16% | 0.34 | 4e-17 | 1e-04 |
|  |  | Lifetime Depression (MHQ) | 1.0 | 99,216 | 0.15 | 1.20% | 0.20 | 7e-228 | 1e-04 |
| PGC MDD  **Incl**. 23andMe | MHQ controls | One Measure | 0.5 | 67,956 | 0.15 | 1.26% | 0.22 | 5e-269 | 1e-04 |
|  |  | Two Measures | 0.4 | 59,001 | 0.15 | 1.96% | 0.27 | 1e-226 | 1e-04 |
|  |  | Three Measures | 0.2 | 36,975 | 0.15 | 2.56% | 0.31 | 9e-159 | 1e-04 |
|  |  | Four Measures | 0.4 | 59,001 | 0.15 | 3.54% | 0.36 | 2e-108 | 1e-04 |
|  |  | Five Measures | 0.1 | 22,831 | 0.15 | 2.62% | 0.37 | 6e-20 | 1e-04 |
|  |  | Lifetime Depression (MHQ) | 1.0 | 99,216 | 0.15 | 1.70% | 0.24 | 2e-237 | 1e-04 |

| **A** | 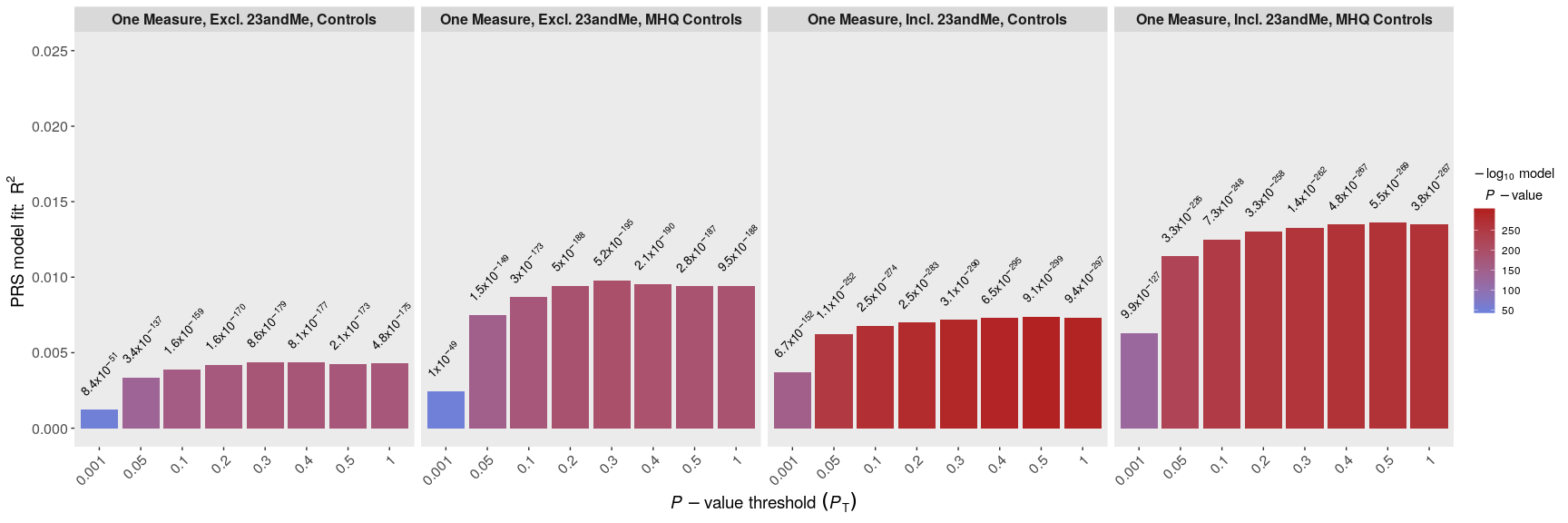 |
| --- | --- |
| **B** | 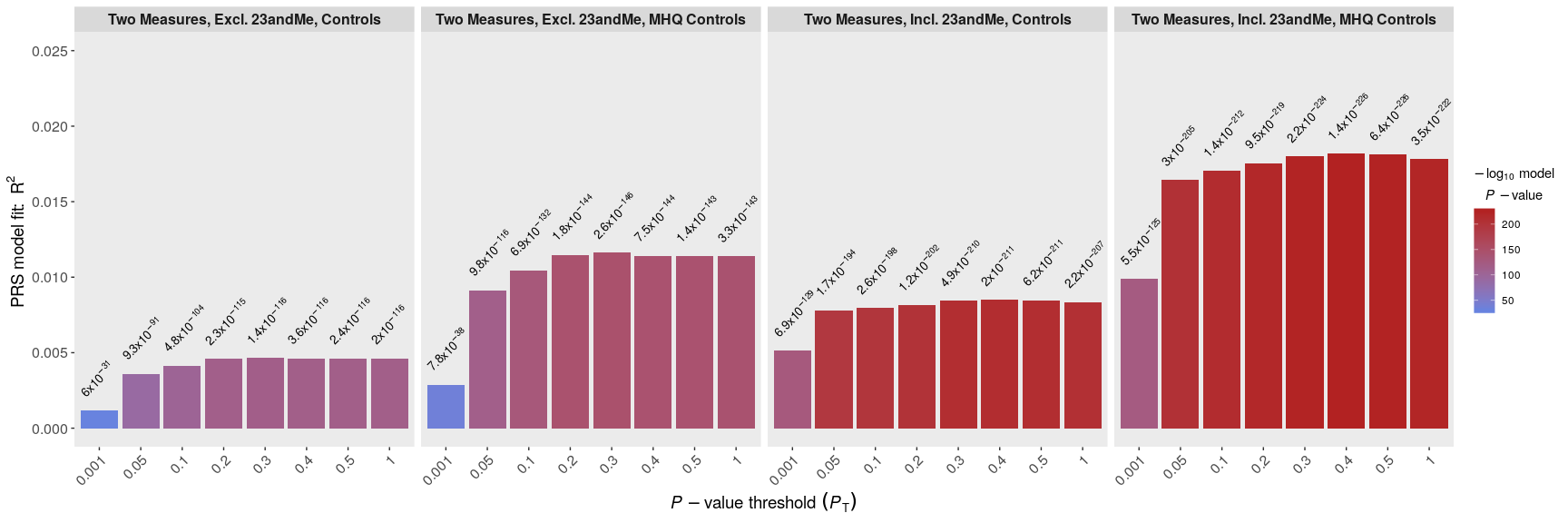 |
| **C** | 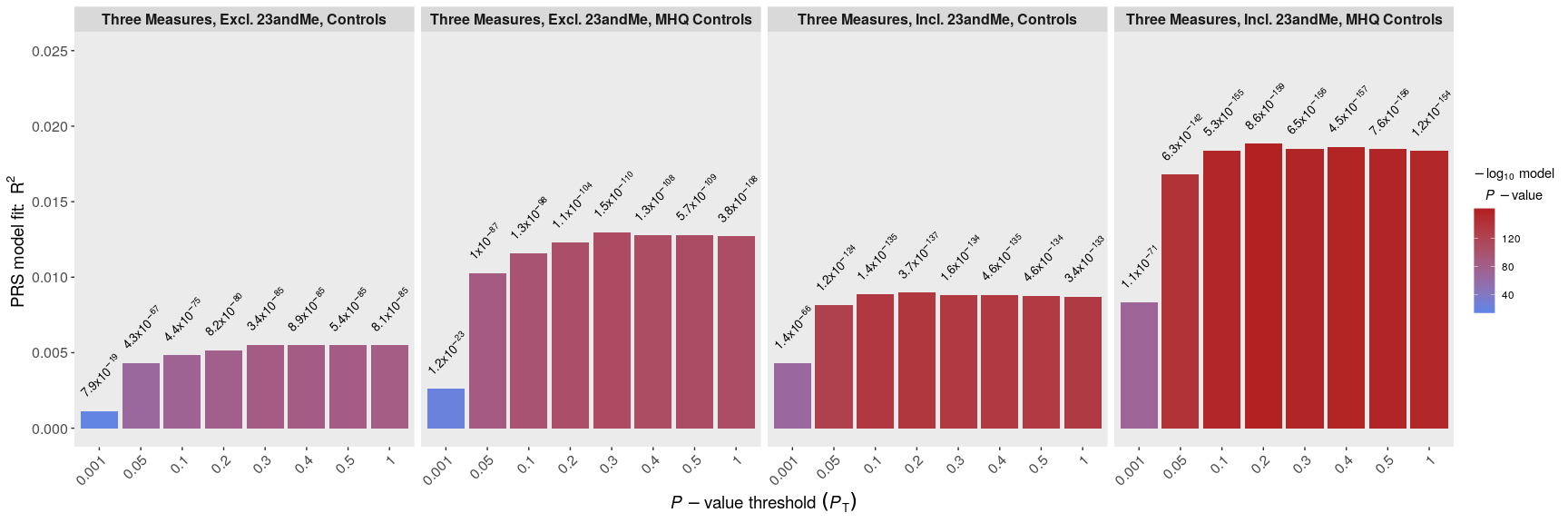 |
| **D** | 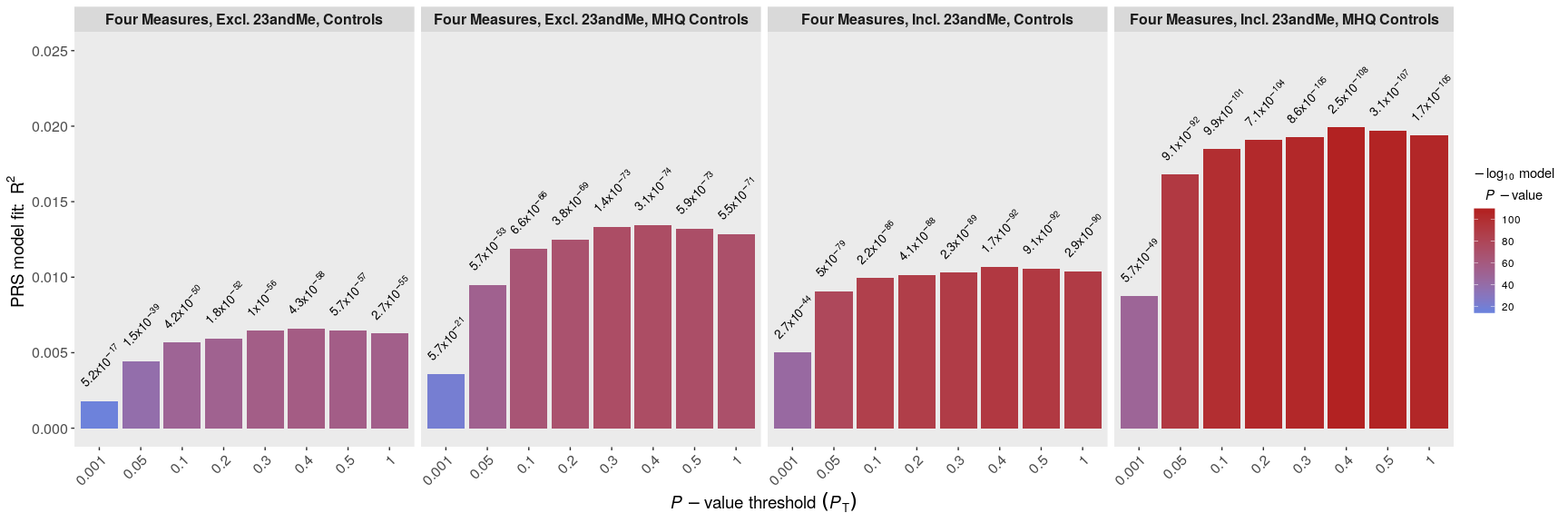 |
| **E** | 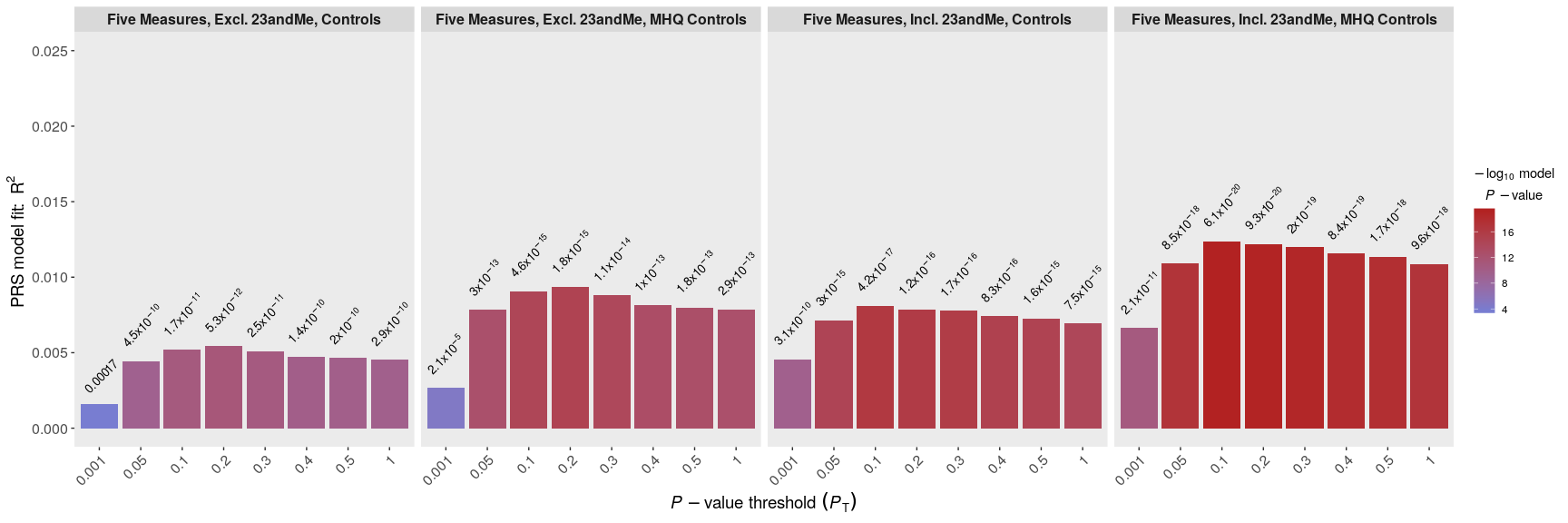 |
| **F** | 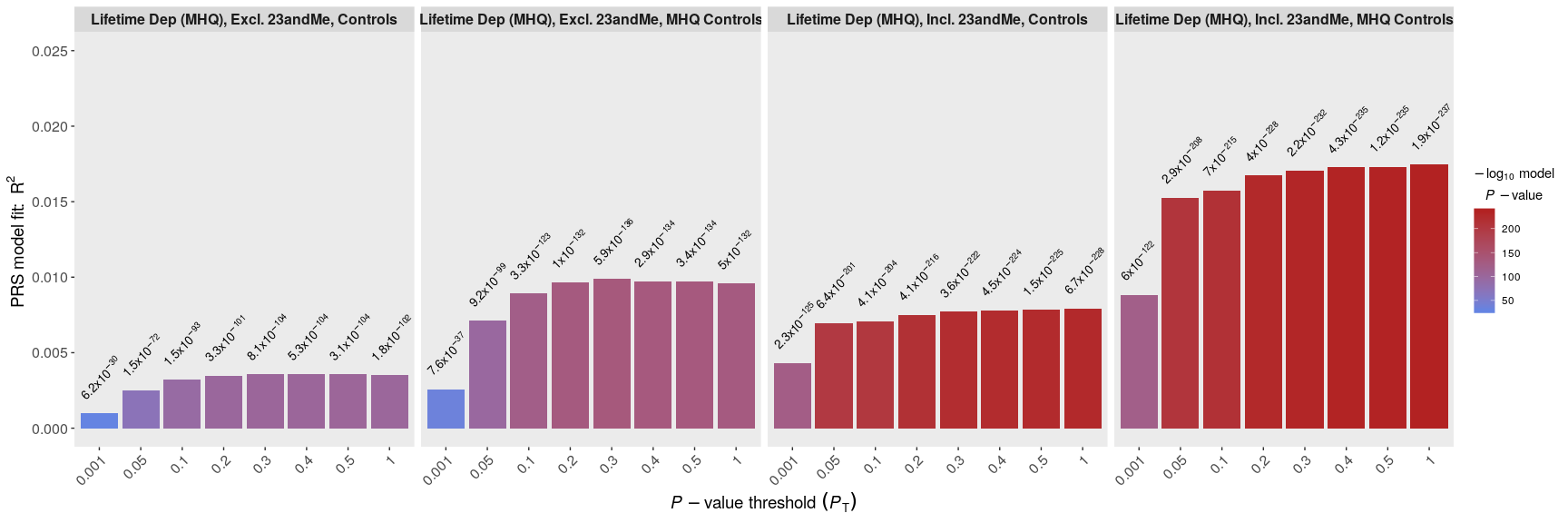 |

**Supplementary Figure 2:** Association between PRS and depression phenotypes across eight *P*_T_. **Row A** = One Measure, **Row B** = Two Measures, **Row C** = Three Measures, **Row D** = Four Measures, **Row E** = Five Measures, **Row F** = Lifetime Depression (MHQ). Excl. 23andMe = PRS calculated using summary statistics from the sub-set of the PGC MDD sample (excluding UKB and 23andMe). Incl. 23andMe = PRS calculated using summary statistics from the full PGC MDD sample (excluding UKB). Observed p-values are shown atop each bar. Y-axis shows unadjusted R^2^ (observed-scale).

### 8 Area Under the Curve Analyses

| **Controls** | | | | |
| --- | --- | --- | --- | --- |
| 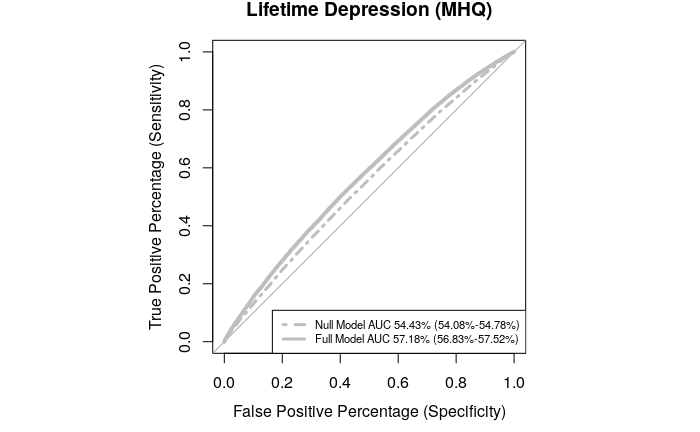 | 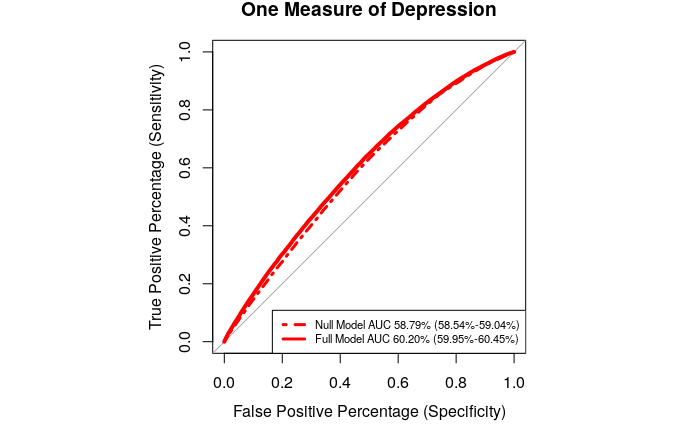 | 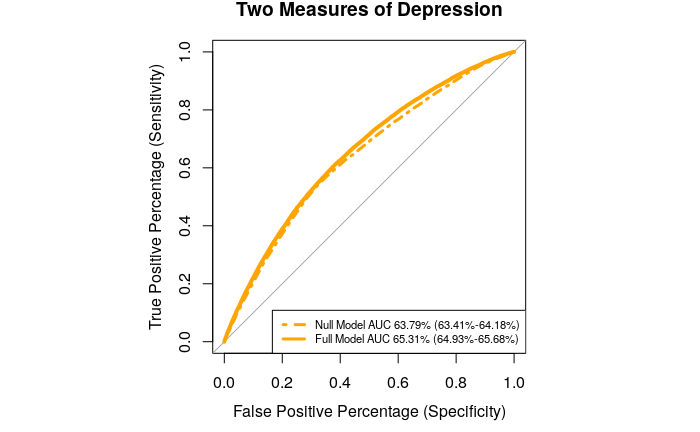 | 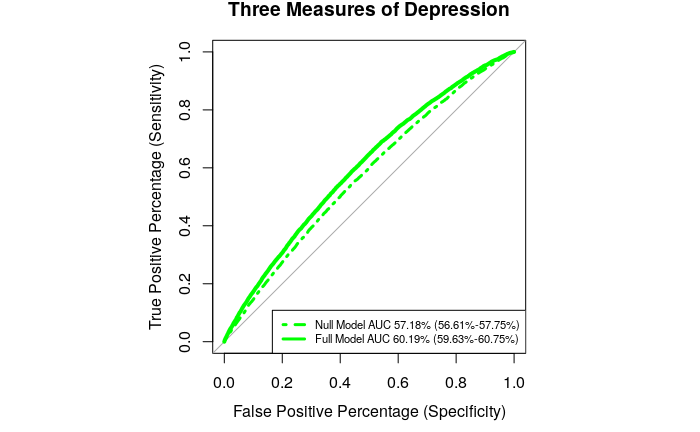 | 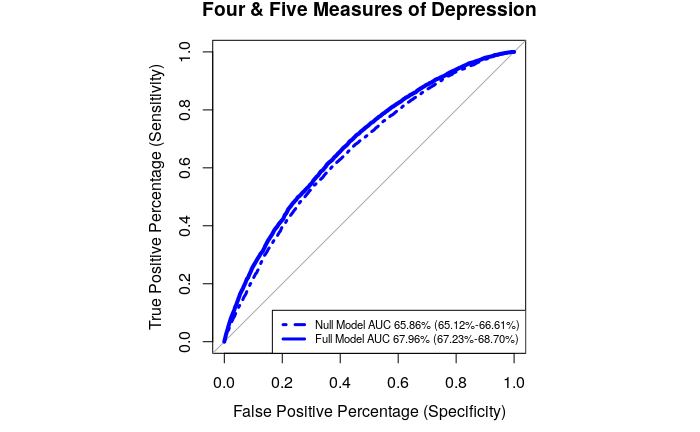 |
| **MHQ controls** | | | | |
| 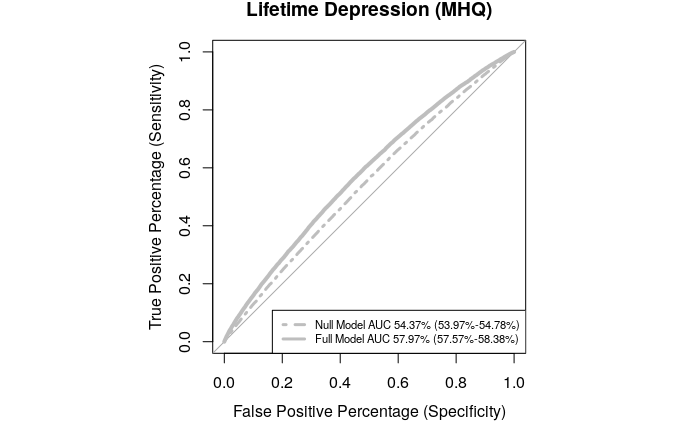 | 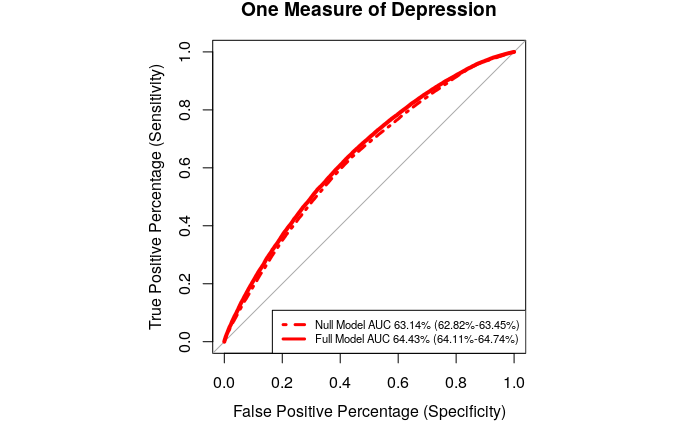 | 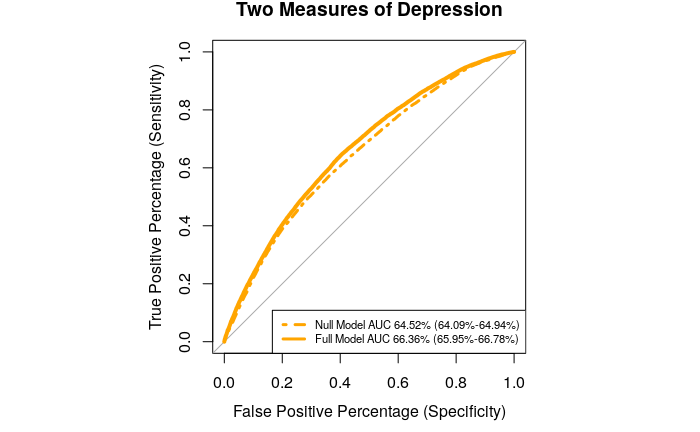 | 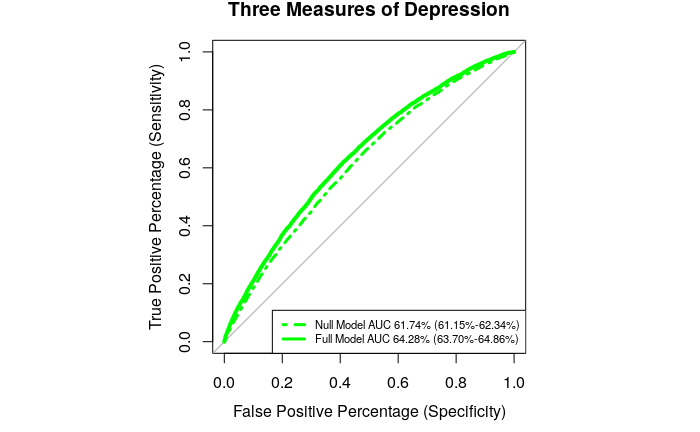 | 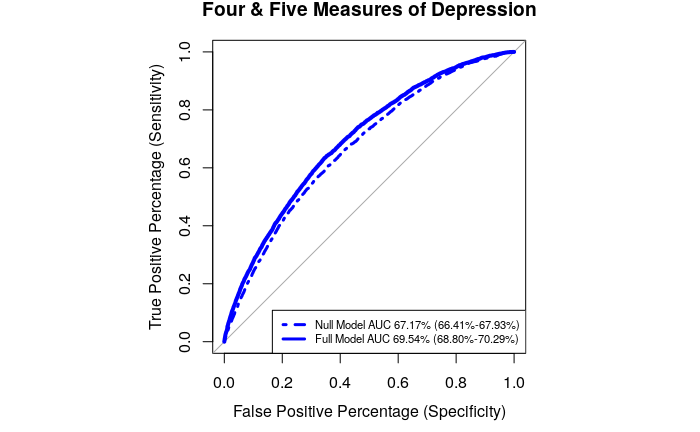 |

**Supplementary Figure 3**: ROC plots. AUC estimates from Null Models (Phenotype ~ 6PCs + Genotyping Batch + Centre of Assessment) and Full Models (Phenotype ~ PRS + 6PCs + Genotyping Batch + Centre of Assessment). PRS calculated using PGC MDD summary statistics including 23andMe at the *P*_T_ identified for each phenotype in PRSice analysis. Logistic regressions performed on depression phenotypes shown in plot titles, using controls (upper panel) and MHQ controls (lower panel).

### 9 GWAS results

| **A** | 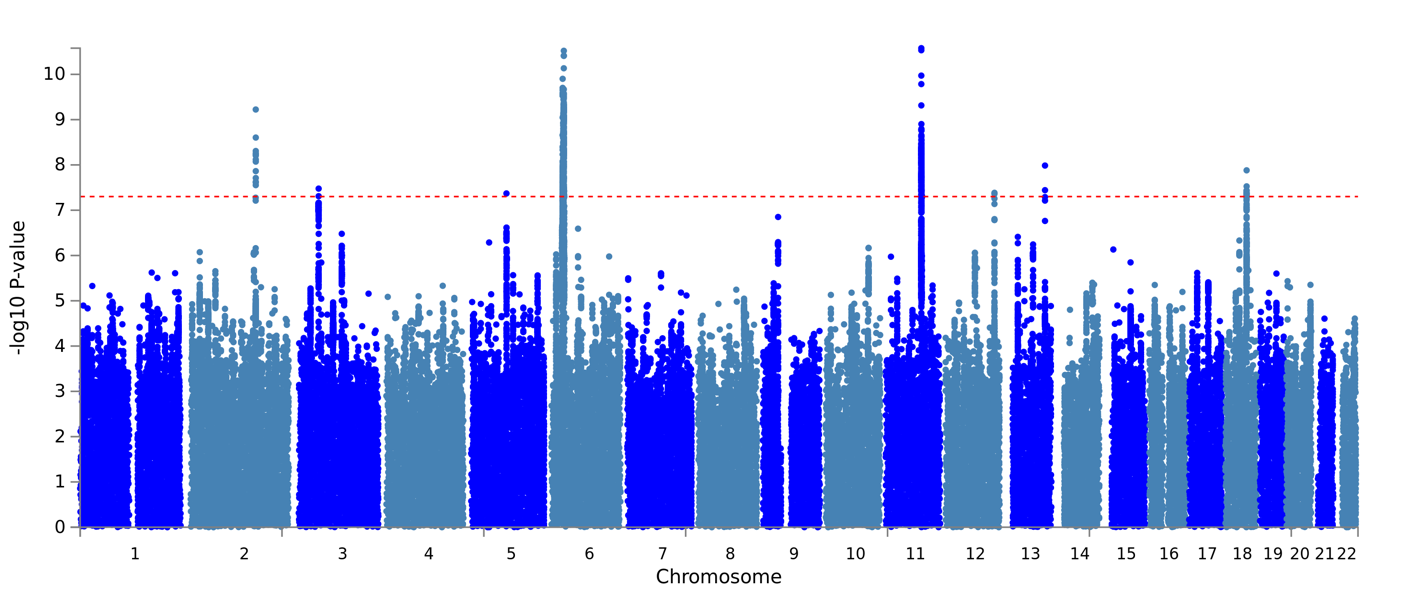 | 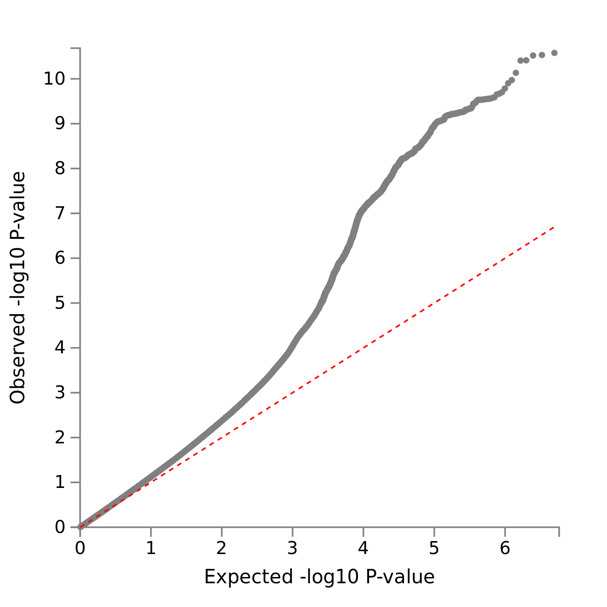 |
| --- | --- | --- |
| **B** | 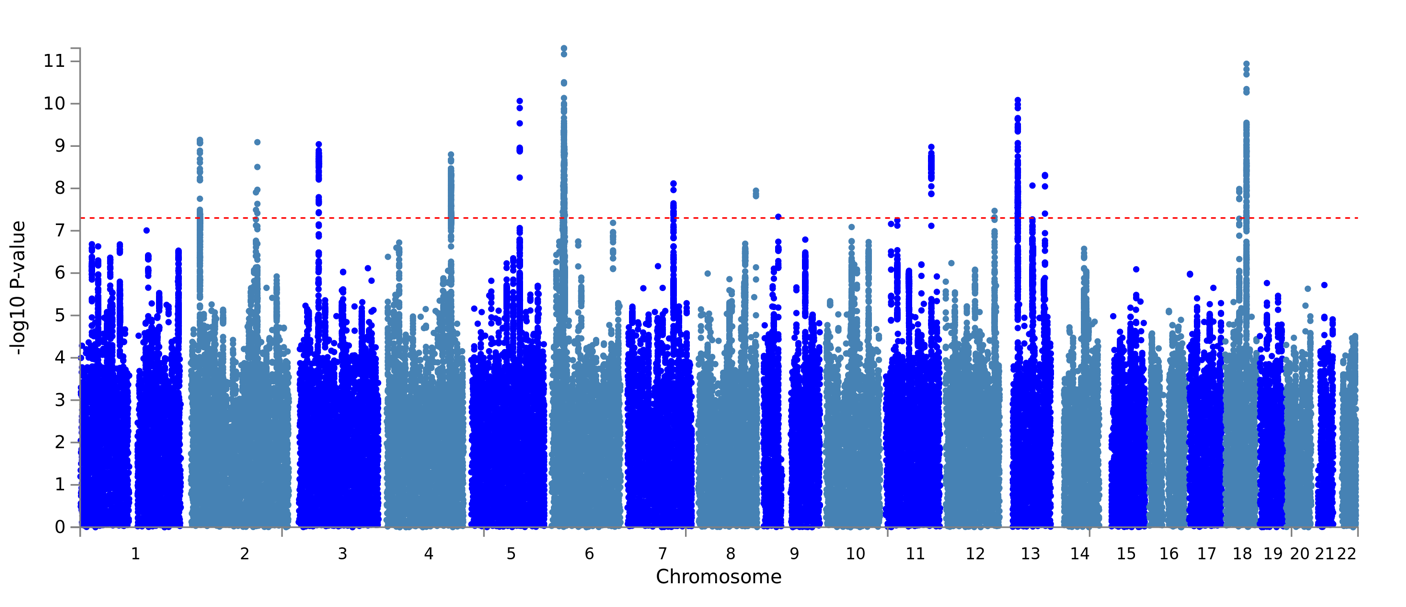 | 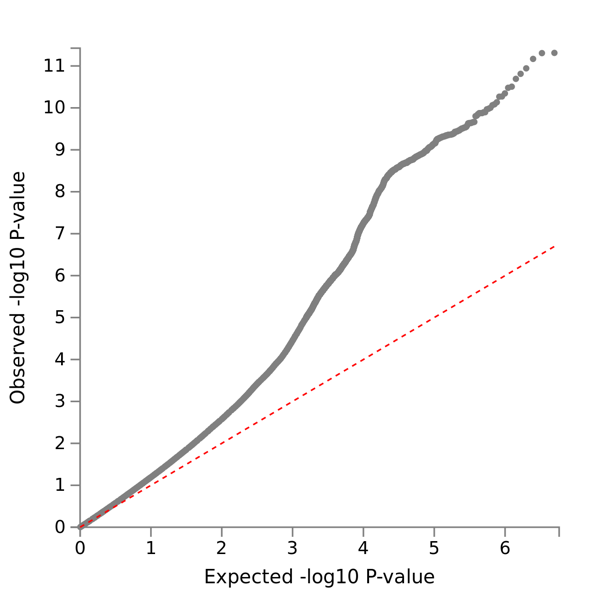 |

**Supplementary Figure 4:** Manhattan and QQ plots from GWAS of One Measure of Depression. **A**: controls, **B**: MHQ controls.

| **A** | 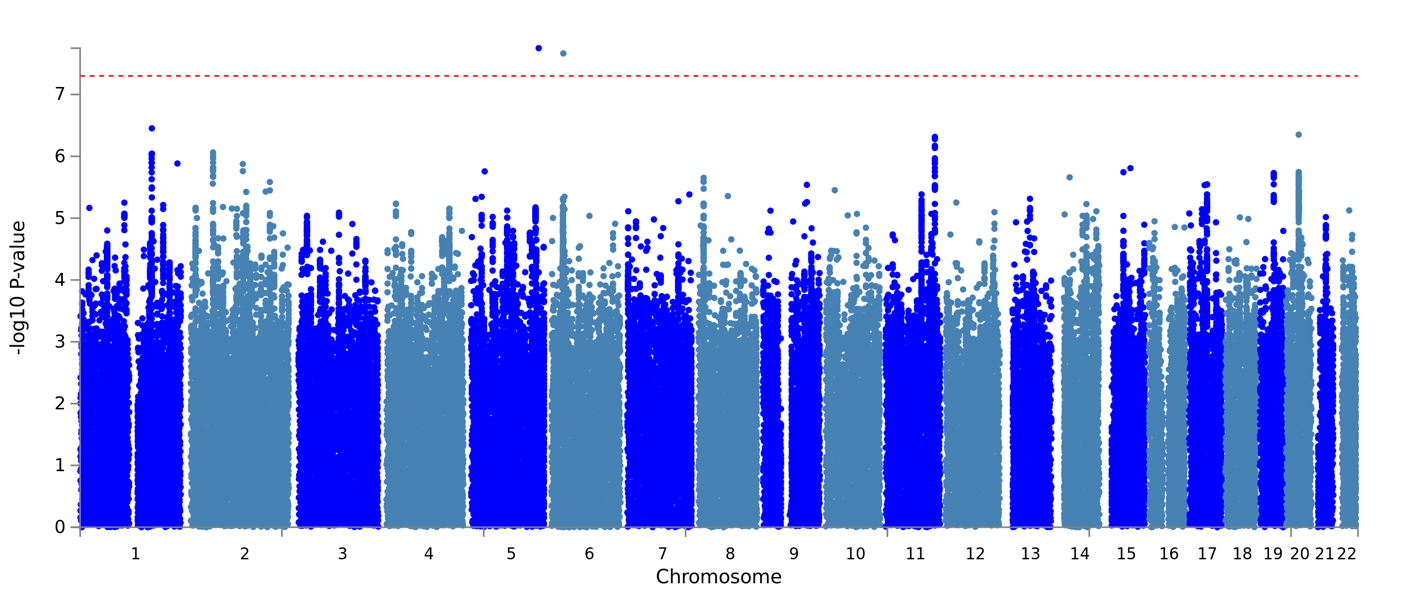 | 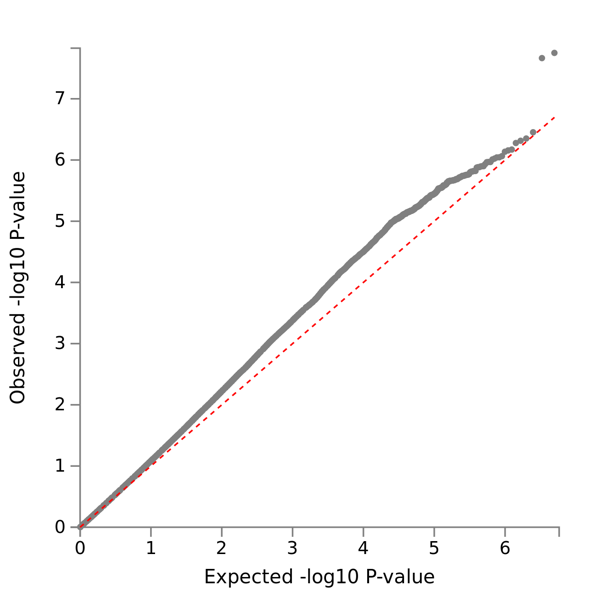 |
| --- | --- | --- |
| **B** | 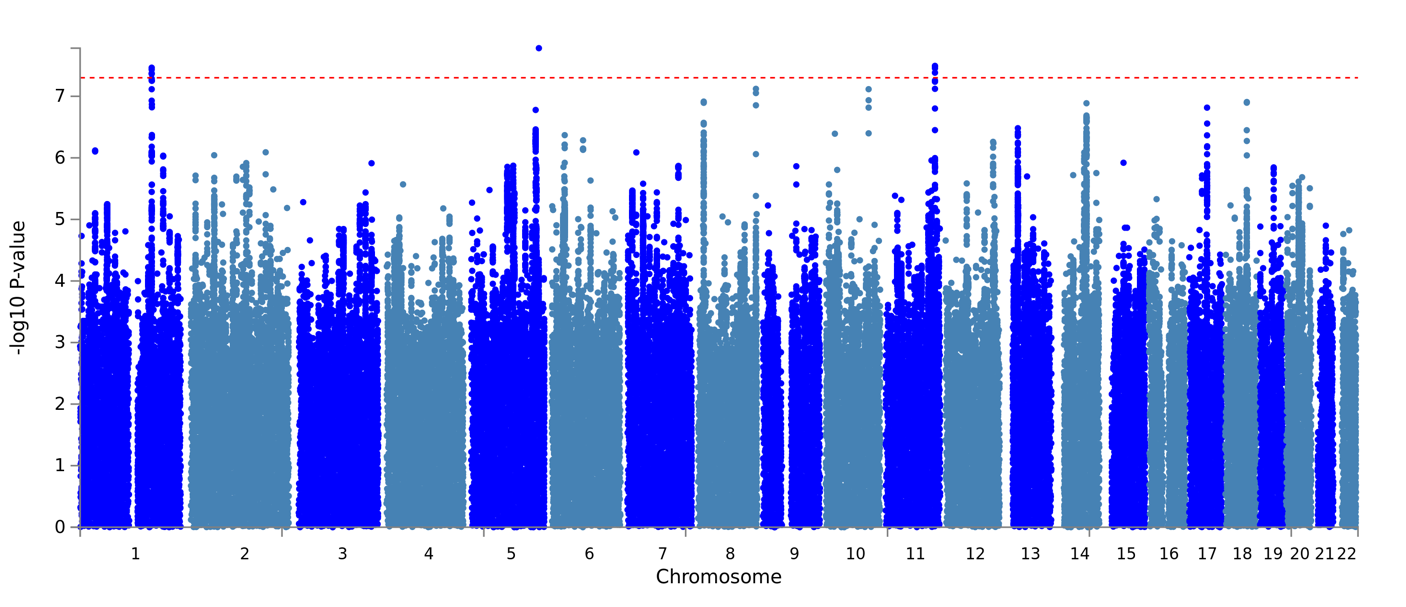 | 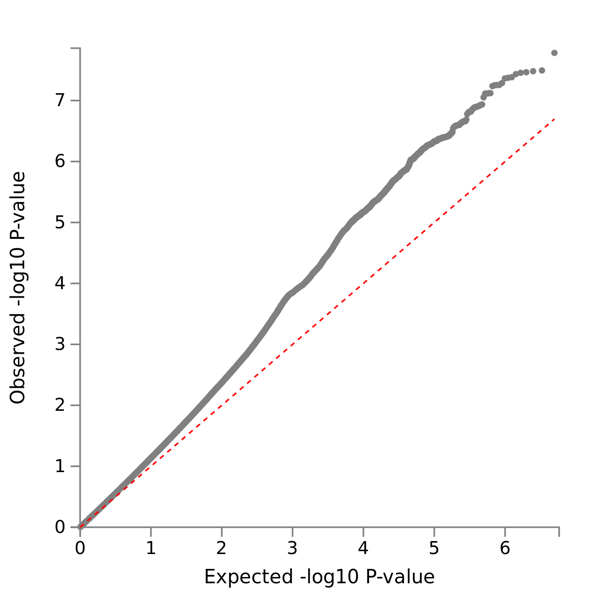 |

**Supplementary Figure 5:** Manhattan and QQ plots from GWAS of Two Measures of Depression. **A**: controls, **B**: MHQ controls.

| **A** | 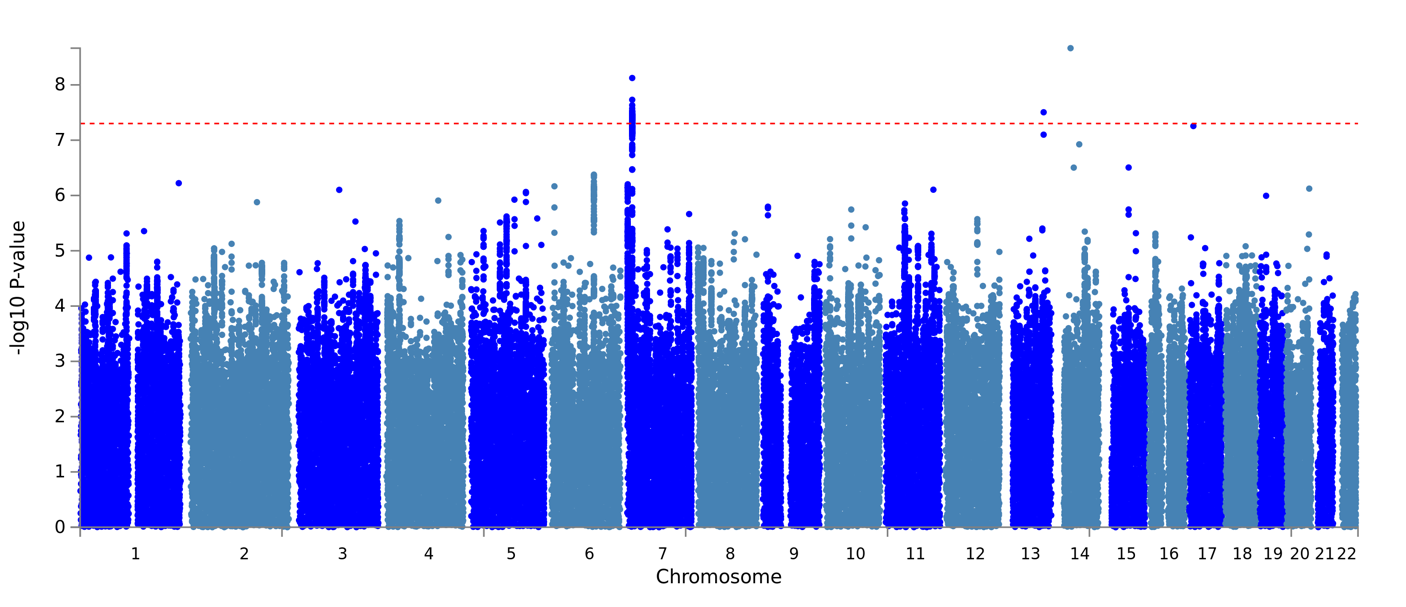 | 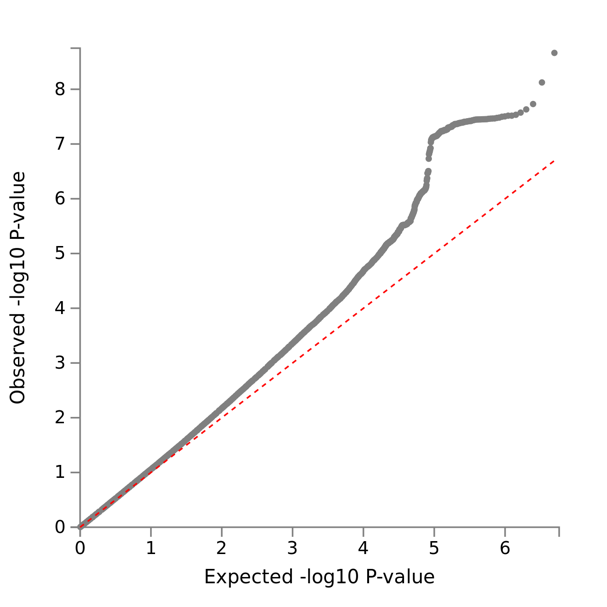 |
| --- | --- | --- |
| **B** | 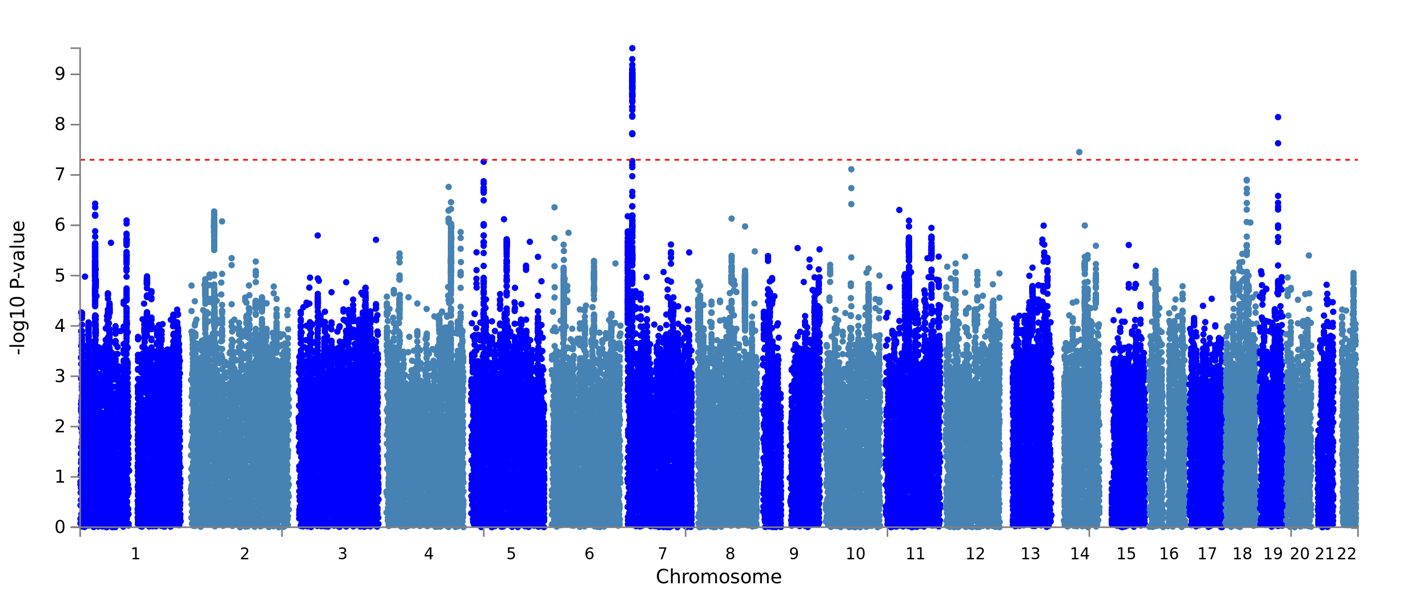 | 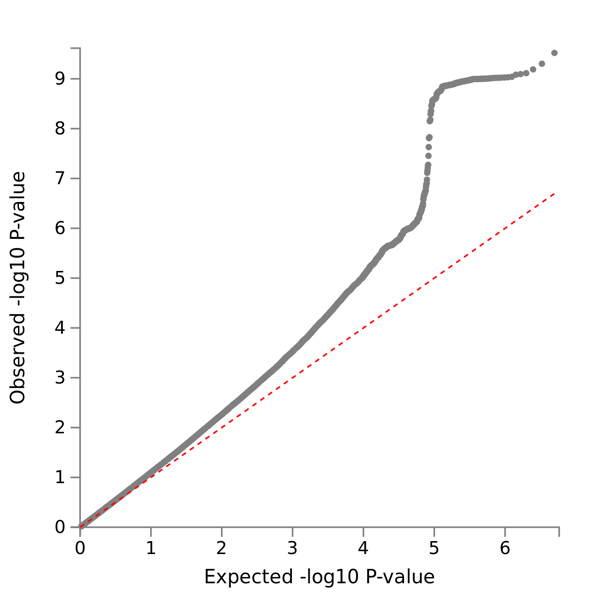 |

**Supplementary Figure 6:** Manhattan and QQ plots from GWAS of Three Measures of Depression. **A**: controls, **B**: MHQ controls.

| **A** | 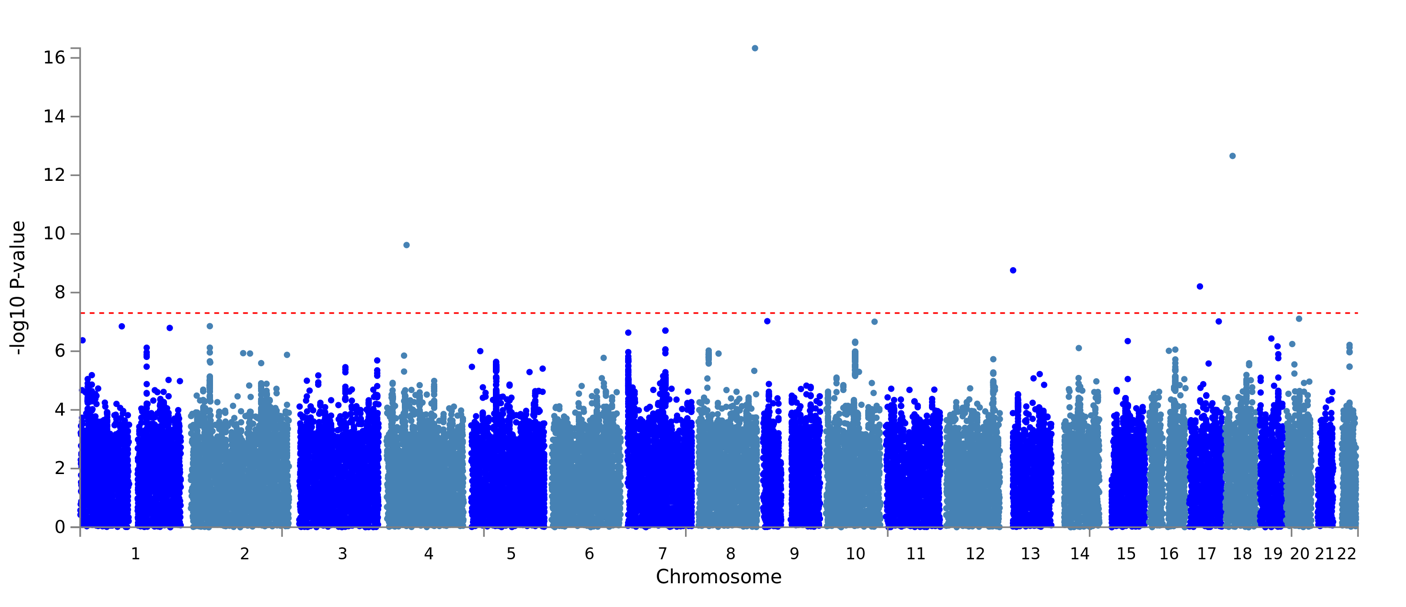 | 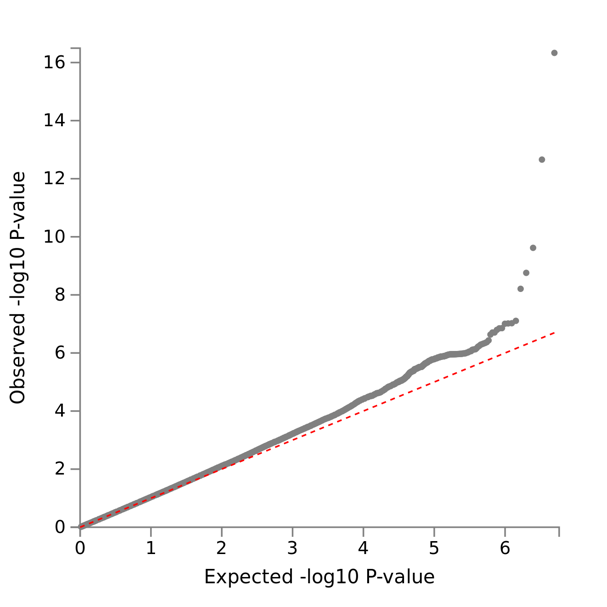 |
| --- | --- | --- |
| **B** | 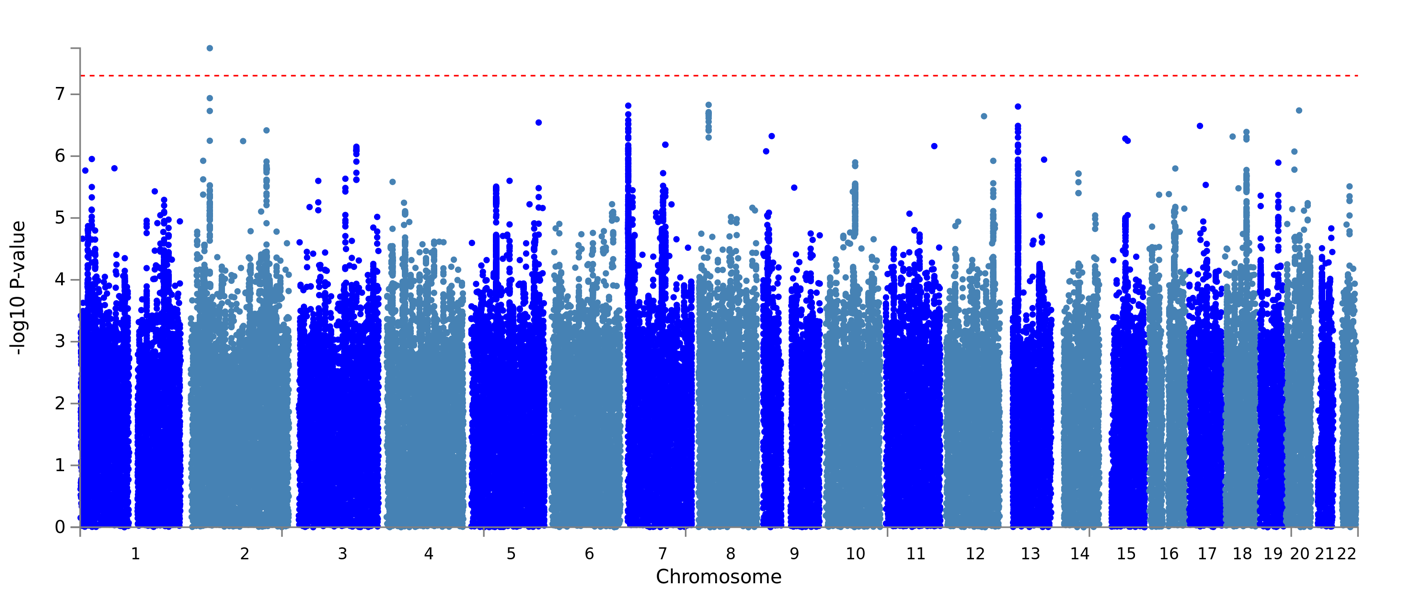 | 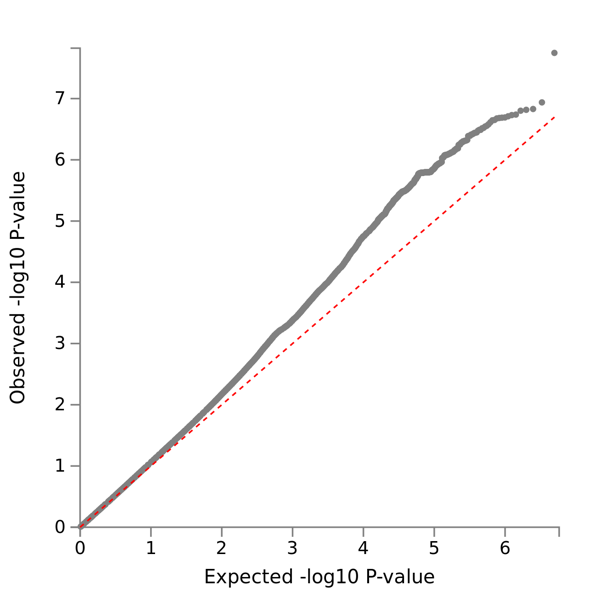 |

**Supplementary Figure 7:** Manhattan and QQ plots from GWAS of Four & Five Measure of Depression. **A**: controls, **B**: MHQ controls.

| **A** | 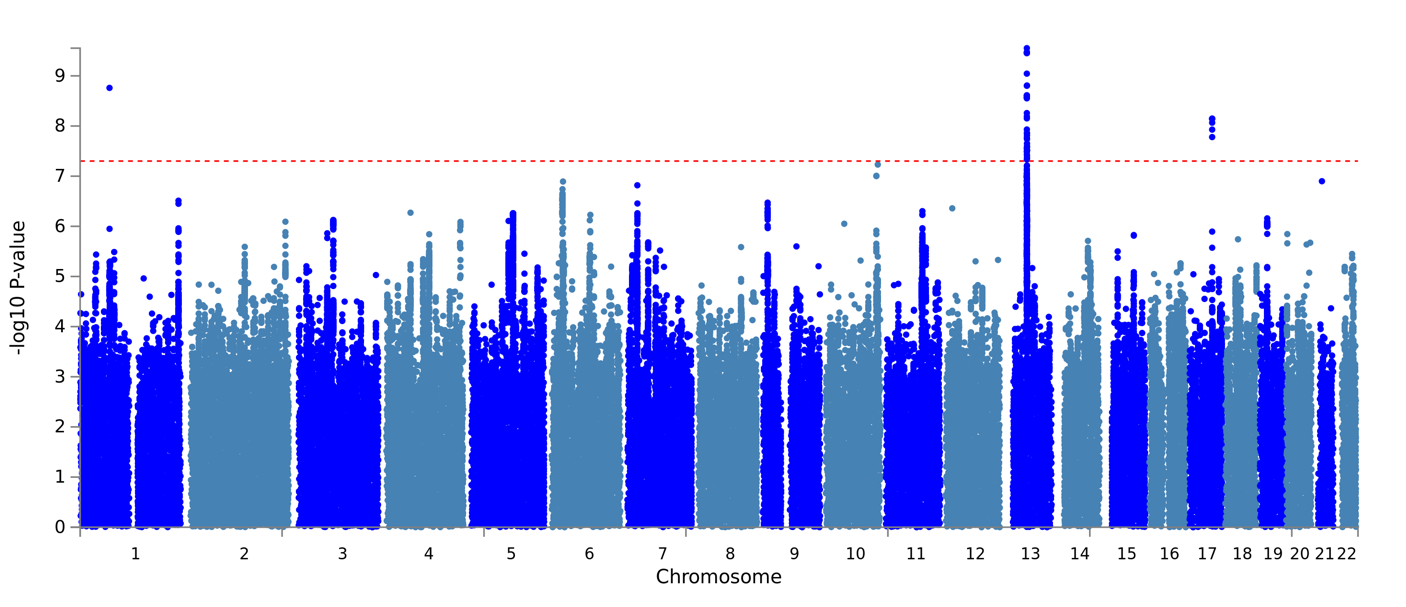 | 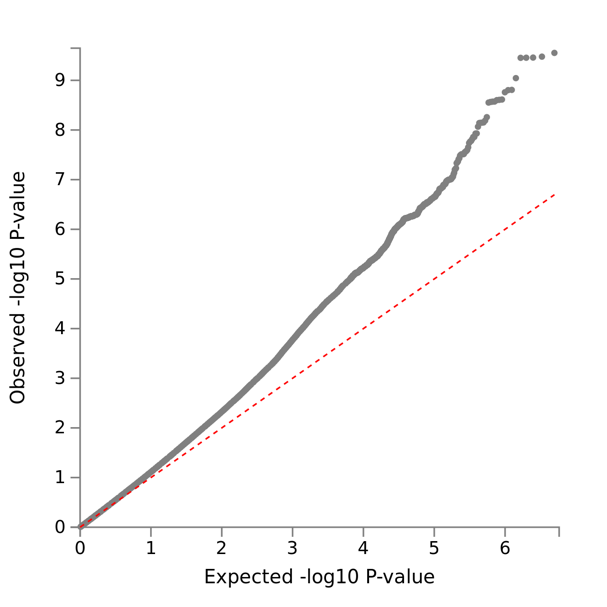 |
| --- | --- | --- |
| **B** | 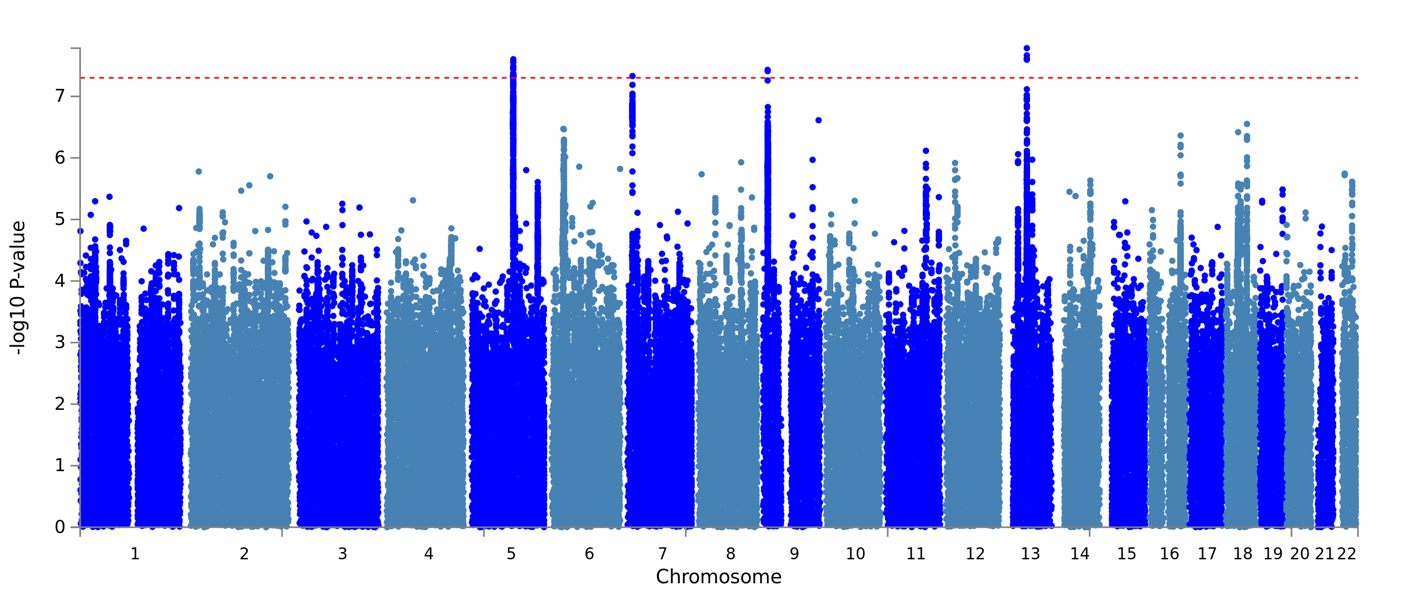 | 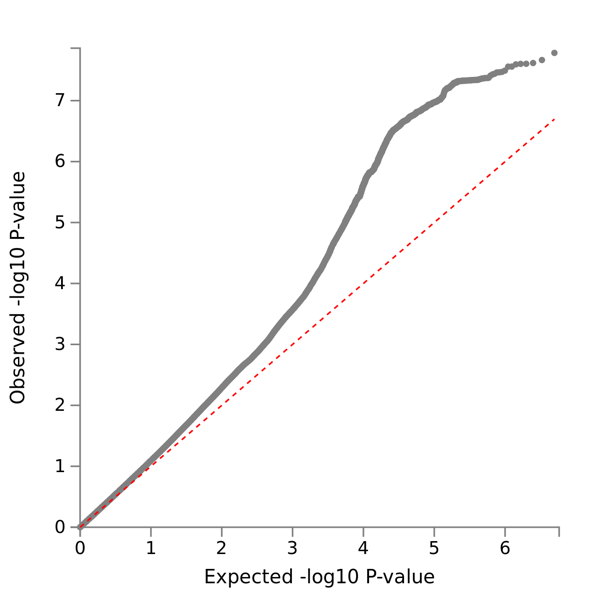 |

**Supplementary Figure 8:** Manhattan and QQ plots from GWAS of Lifetime Depression (MHQ). **A**: controls, **B**: MHQ controls.

GWAS results have been published on FUMA^4^ (<https://fuma.ctglab.nl/>) to show details of significant SNPs including evidence for association with other traits in the GWAS catalogue.

**Supplementary Table 13**: FUMA titles and IDs for GWAS performed in this study.

| **GWAS Trait** | **Controls** | **FUMA ID** | **FUMA Title** |
| --- | --- | --- | --- |
| One Measure | Controls | 151 | One_232k_controls |
| Two Measures |  | 153 | Two_232k_controls |
| Three Measures |  | 152 | Three_232k_controls |
| Four Measures |  | 154 | Four_five_232k_controls |
| Lifetime Depression (MHQ) |  | 150 | MHQ_232k_controls |
| One Measure | MHQ controls | 146 | One_58k_controls |
| Two Measures |  | 147 | Two_58k_controls |
| Three Measures |  | 148 | Three_58k_controls |
| Four & Five Measures |  | 149 | Four_five_58k_controls |
| Lifetime Depression (MHQ) |  | 145 | MHQ_58k_controls |

**Supplementary Table 14:** GWAS and *h*^2^_SNP_ results: depression cases compared to **controls**.

|  | **Depression phenotype in the UKB** | | | | |
| --- | --- | --- | --- | --- | --- |
| **Parameter** | **Lifetime Depression (MHQ)** | **One Measure** | **Two Measures** | **Three Measures** | **Four & Five Measures** |
| **GWAS results (BGENIE)** | | | | | |
| No. SNPs in GWAS | 9,940,917 | 9,940,917 | 9,940,917 | 9,940,917 | 9,940,917 |
| Mean chi^2^ | 1.162 | 1.182 | 1.115 | 1.083 | 1.053 |
| Lambda GC | 1.138 | 1.151 | 1.103 | 1.076 | 1.045 |
| Max chi^2^ | 39.807 | 44.425 | 31.343 | 33.399 | 70.477 |
| No. Significant SNPs | 40 | 692 | 1 | 67 | 5 |
| **Heritability results (LDSC)** | | | | | |
| Population Prevalence | 0.15 | 0.15 | 0.15 | 0.15 | 0.15 |
| Sample Prevalence | 0.110 | 0.197 | 0.084 | 0.040 | 0.020 |
| No. SNPs post-munge | 7,353,982 | 7,353,982 | 7,353,982 | 7,353,982 | 7,353,982 |
| No. SNPs post LD merge | 1,184,555 | 1,184,555 | 1,184,555 | 1,184,555 | 1,184,555 |
| Liability scale h^2^ (SE) | 0.1134 (0.008) | 0.0723 (0.0047) | 0.1121 (0.0093) | 0.1874 (0.0183) | 0.2086 (0.0289) |
| Liability scale h^2^ 95% CIs | 0.098 - 0.129 | 0.063 - 0.082 | 0.094 - 0.130 | 0.152 - 0.223 | 0.152 - 0.265 |
| Lambda | 1.1619 | 1.194 | 1.127 | 1.0988 | 1.0557 |
| Mean Chi^2^ | 1.1949 | 1.2186 | 1.1379 | 1.1058 | 1.0599 |
| Intercept (SE) | 1.0047 (0.0076) | 1. (0.0076) | 0.9954 (0.0068) | 0.9923 (0.0069) | 0.9971 (0.0068) |

**Supplementary Table 15:** GWAS and *h*^2^_SNP_ results: depression cases compared to **MHQ** **controls**.

|  | **Depression phenotype in the UKB** | | | | |
| --- | --- | --- | --- | --- | --- |
| **Parameter** | **Lifetime Depression (MHQ)** | **One Measure** | **Two Measures** | **Three Measures** | **Four & Five Measures** |
| **GWAS results (BGENIE)** | | | | | |
| No. SNPs in GWAS | 9,940,917 | 9,940,917 | 9,940,917 | 9,940,917 | 9,940,917 |
| Mean chi^2^ | 1.137 | 1.282 | 1.193 | 1.137 | 1.086 |
| Lambda GC | 1.12 | 1.226 | 1.171 | 1.125 | 1.077 |
| Max chi^2^ | 31.349 | 47.732 | 30.521 | 39.661 | 31.705 |
| No. Significant SNPs | 52 | 786 | 8 | 85 | 1 |
| **Heritability results (LDSC)** | | | | | |
| Population Prevalence | 0.15 | 0.15 | 0.15 | 0.15 | 0.15 |
| Sample Prevalence | 0.333 | 0.497 | 0.270 | 0.144 | 0.077 |
| No. SNPs post-munge | 7,353,982 | 7,353,982 | 7,353,982 | 7,353,982 | 7,353,982 |
| No. SNPs post LD merge | 1,184,555 | 1,184,555 | 1,184,555 | 1,184,555 | 1,184,555 |
| Liability scale h^2^ (SE) | 0.1268 (0.0091) | 0.1729 (0.0091) | 0.2259 (0.0132) | 0.3315 (0.0212) | 0.336 (0.0337) |
| Liability scale h^2^ 95% CIs | 0.109 - 0.145 | 0.155 - 0.191 | 0.200 - 0.252 | 0.290 - 0.373 | 0.270 - 0.402 |
| Lambda | 1.1428 | 1.2831 | 1.207 | 1.1587 | 1.0895 |
| Mean Chi^2^ | 1.1599 | 1.3391 | 1.2293 | 1.1697 | 1.0966 |
| Intercept (SE) | 0.9984 (0.0071) | 1.0136 (0.0085) | 1.0014 (0.0074) | 0.9911 (0.0072) | 0.9998 (0.007) |

### 10 Genetic Correlations

**Supplementary Table 16:** Genetic correlations between UKB and PGC depression phenotypes. PGC Incl. 23andMe = summary statistics from the full PGC MDD sample (excluding UKB). PGC excl. 23andMe = summary statistics from the sub-set of the PGC MDD sample with self-reported cases removed (excluding UKB and 23andMe). Pairwise *r*_G_ estimates were calculated using summary statistics generated from GWASs of UKB depression phenotypes using controls and MHQ controls.

| **Phenotype 1** | **Phenotype 2** | ***r*_G_** | **95% CIs** | **P-value** | ***r*_G_** | **95% CIs** | **P-value** |
| --- | --- | --- | --- | --- | --- | --- | --- |
|  | | **Controls** | | | **MHQ controls** | | |
| Lifetime Depression (MHQ) | One Measure | 0.53 | 0.46 - 0.60 | 3e-48 | 0.71 | 0.66 - 0.76 | 3e-152 |
| Lifetime Depression (MHQ) | Two Measures | 0.62 | 0.53 - 0.71 | 1e-43 | 0.79 | 0.73 - 0.85 | 7e-138 |
| Lifetime Depression (MHQ) | Three Measures | 0.52 | 0.42 - 0.62 | 4e-26 | 0.76 | 0.68 - 0.84 | 2e-85 |
| Lifetime Depression (MHQ) | Four and Five Measures | 0.50 | 0.36 - 0.64 | 1e-12 | 0.73 | 0.62 - 0.84 | 1e-37 |
| Lifetime Depression (MHQ) | PGC Incl. 23andMe | 0.80 | 0.74 - 0.86 | 2e-140 | 0.88 | 0.80 - 0.96 | 1e-112 |
| Lifetime Depression (MHQ) | PGC excl. 23andMe | 0.72 | 0.63 - 0.81 | 9e-57 | 0.90 | 0.80 - 1.00 | 2e-65 |
| One Measure | Two Measures | 1.00 | 0.92 - 1.08 | 6e-118 | 0.99 | 0.95 - 1.03 | 0e+00 |
| One Measure | Three Measures | 0.90 | 0.81 - 0.99 | 2e-76 | 0.95 | 0.90 - 1.00 | 1e-260 |
| One Measure | Four and Five Measures | 0.94 | *0.79 - 1.09 | 9e-33 | 0.96 | *0.87 - 1.05 | 2e-93 |
| One Measure | PGC Incl. 23andMe | 0.84 | 0.78 - 0.90 | 5e-167 | 0.65 | 0.60 - 0.70 | 5e-126 |
| One Measure | PGC excl. 23andMe | 0.86 | 0.77 - 0.95 | 3e-79 | 0.74 | 0.66 - 0.82 | 2e-78 |
| Two Measures | Three Measures | 0.86 | 0.74 - 0.98 | 1e-43 | 0.92 | 0.85 - 0.99 | 5e-158 |
| Two Measures | Four and Five Measures | 0.88 | 0.70 - 1.06 | 1e-21 | 0.94 | 0.83 - 1.05 | 3e-64 |
| Two Measures | PGC Incl. 23andMe | 0.82 | 0.75 - 0.89 | 1e-106 | 0.67 | 0.61 - 0.73 | 1e-110 |
| Two Measures | PGC excl. 23andMe | 0.86 | 0.76 - 0.96 | 6e-68 | 0.75 | 0.67 - 0.83 | 1e-81 |
| Three Measures | Four and Five Measures | *1.01 | 0.81 - 1.21 | 2e-22 | *1.01 | 0.89 - 1.13 | 6e-61 |
| Three Measures | PGC Incl. 23andMe | 0.72 | 0.64 - 0.80 | 7e-76 | 0.62 | 0.57 - 0.67 | 1e-107 |
| Three Measures | PGC excl. 23andMe | 0.76 | 0.64 - 0.88 | 4e-37 | 0.69 | 0.60 - 0.78 | 4e-50 |
| Four and Five Measures | PGC Incl. 23andMe | 0.78 | 0.65 - 0.91 | 6e-34 | 0.69 | 0.60 - 0.78 | 1e-50 |
| Four and Five Measures | PGC excl. 23andMe | 0.86 | *0.70 - 1.02 | 6e-25 | 0.81 | 0.69 - 0.93 | 2e-39 |
| PGC Incl. 23andMe | PGC excl. 23andMe | 0.97 | 0.94 - 1.00 | 0e+00 | 0.97 | 0.94 - 1.00 | 0e+00 |

*LDSC is not a bounded estimator and can produce estimates outside the range or -1, 1 when standard errors are large due to small samples. This issue is discussed in the LDSC GitHub issue log: <https://github.com/bulik/ldsc/issues/89>

| **A) Controls**  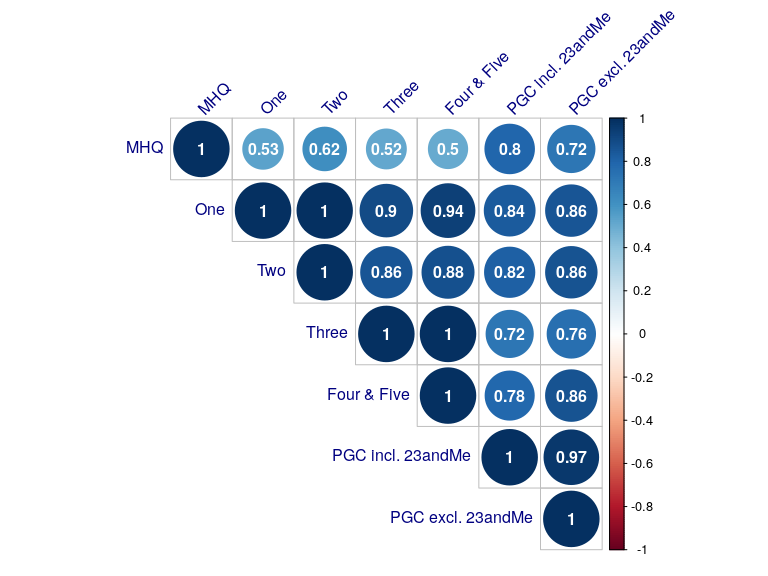 | **B) MHQ controls**  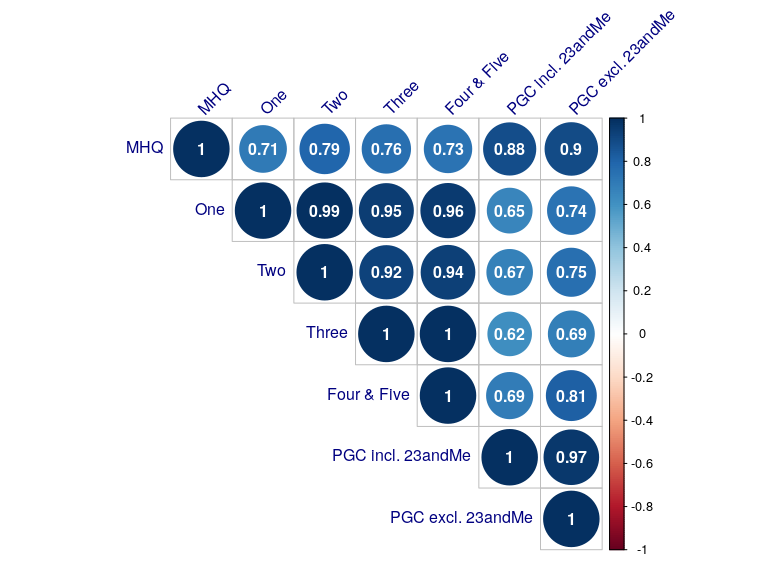 |
| --- | --- |

**Supplementary Figure 9:** Genetic correlation matrix. Circle sizes correspond to the magnitude of correlation, increasing as *r*_G_ approaches 1. MHQ = Lifetime Depression (MHQ). One, Two, Three, Four & Five = cases determined by number of observed depression measures. PGC Incl. 23andMe = summary statistics from the full PGC MDD sample (excluding UKB). PGC excl. 23andMe = summary statistics from the sub-set of the PGC MDD sample with self-reported cases removed (excluding UKB and 23andMe). Summary statistics used to estimate *r*_G_ were generated from GWASs of UKB depression phenotypes using controls (left matrix) and MHQ controls (right matrix).
